# Assessing Inter-platform Variability of Blood Parameters across Three Automated Platforms: Report from Multi-centric Phenome India Study

**DOI:** 10.64898/2026.01.24.26344729

**Authors:** Md. Intyaz Ali, Mamta Rathore, Rajat Ujjainiya, Satyartha Prakash, Ankita Sahu, Shilpa Ray, Nancy Rawat, Rakesh Kanakrajan Vijyakumari, Manuj Kr Das, Milind Sanjay Kale, Saheli Chowdhury, Partitusti Basu, Aastha Mishra, Kumardeep Chaudhary, Swasti Raychaudhuri, Karthik Bharadwaj Tallapaka, Partha Chakraborty, Umakanta Subudhi, Mahesh J Kulkarni, Jatin Kalita, Ashish Awasthi, Phenome India Consortium, Viren Sardana, Shantanu Sengupta

## Abstract

**Background:** Accurate and consistent laboratory measurements are essential for reliable diagnosis and clinical decision making. However, variability among automated analyzers can compromise diagnostic reliability and in population studies, instrument-specific differences in estimates hinder meaningful cross-study comparisons.

**Objective:** To evaluate inter-platform variability and interchangeability of laboratory parameters relevant to disease diagnosis across Beckman, Siemens and Roche automated analyzers.

**Methods:** From multi-centric Phenome India study, 400 blood samples were analysed for 29 parameters (25 clinical chemistry and 4 immunological) on the three platforms. Inter-platform comparability was assessed using Passing-Bablok regression, Bland-Altman analysis and concordance correlation coefficient (CCC) to evaluate biases and overall agreement. Platform-specific thresholds for low Vitamin B12 were estimated via multivariable regression, adjusting for Folate relative to homocysteine.

**Results:** Inter-platform variability differed across parameters. Enzymatic markers (ALT, AST, ALP, GGT), Urea, Uric acid and HbA1c showed excellent agreement (CCC >0.96). Lipid assays showed mixed concordance: Total cholesterol and Triglycerides correlated strongly, whereas LDL-C and apolipoproteins (ApoA1, ApoB) exhibited systematic bias. Direct Bilirubin also showed divergence. Among immunoassays, Ferritin, Vitamin D and TSH had moderate to strong agreement, but Vitamin B12 showed poor agreement, with low cut-offs, adjusted for Folate and defined by the homocysteine inflection point, at 100 pg/mL (Beckman), 280 pg/mL (Siemens) and 330 pg/mL (Roche).

**Conclusion:** Despite consistent results within platforms, inter-platform agreement varies widely by parameter. This underscores the need for local validation, instrument-specific reference intervals and harmonisation to ensure diagnostic accuracy and enable meaningful cross-study comparisons.

## Introduction

Laboratory testing underpins modern medical practice by providing essential data for patient diagnosis, monitoring and treatment [1]. Accuracy and consistency in clinical chemistry results are critical since even minor deviations can influence clinical decisions, potentially leading to misdiagnosis, inappropriate treatment or delayed intervention [2]. However, variability among automated analyzers, arising from differences in instrument design, calibration methods, environmental conditions, reagent composition and maintenance, remains a persistent challenge. Such variability is particularly relevant for clinical chemistry and immunoassay parameters including electrolytes, lipids and proteins, which are key markers in metabolic and cardiovascular diseases [3–5]. Although efforts towards standardisation continue globally, true result interchangeability across platforms is limited, complicating both clinical diagnostics and population-based research. Manufacturers calibrate analyzers using proprietary methods and reference materials, introducing systematic biases even when testing identical samples. This lack of harmonisation complicates not only clinical use but also large-scale studies that rely on pooled or comparative data from multiple laboratories. Differing assay characteristics produce inconsistent reference ranges and diagnostic cut-offs. For example, a low Vitamin B12 level defined by one instrument may not correspond to the same clinical threshold on another instrument, posing challenges for inter-study comparisons and meta-analyses. Hence, systematic evaluation of inter-analyzers comparability is essential to improve diagnostic accuracy, data harmonisation and reliability of clinical and epidemiological research. Previous studies have compared commonly used clinical analyzers such as Roche Cobas, Siemens Dimension/Atellica, Abbott Architect and Beckman systems, reporting overall correlations as well as systematic biases and assay-specific variations in reference intervals. For instance, Ruffling et al. compared the analytical performance of the Roche Cobas 6000 and Atellica CI analyzers for 17 analytes (13 chemistry and 4 immunochemistry) using remnant patient samples and observed constant bias in 10 parameters, including Uric acid, amylase, TSH and total HCG [6]. Duz et al. compared the analytical performance parameters, including total analytical error, bias and measurement uncertainty, for 15 biochemistry parameters using Beckman and Cobas analyzers and observed similar outcomes between the two devices. However, over the study period, nearly all tests failed to meet the EuBIVAS, RCPA and percentage permissible uncertainty (pU%) criteria [7]. Similarly, Song et al., at a China-based hospital laboratory, compared the analytical performance of Beckman and Siemens analyzers for routine clinical chemistry tests, including ions, blood lipids, kidney function, liver function and the myocardial enzyme spectrum. They reported significant differences in ALT, Urea, and Total Protein measurements [3]. Ferraro et al. reported inter-method bias for the measurement of total and free prostate-specific antigen across Roche, Beckman, Siemens and Abbott analyzers, highlighting the risk of missing advanced prostate cancer if appropriate conversion factors are not applied [8]. However, these studies mostly focused on limited analytes, small sample sizes and single-center settings, thereby limiting generalizability. The present study addresses gaps by conducting a large-scale, multi-centric evaluation using field-level sample collection and a broader panel of 29 clinical chemistry and immunoassay parameters. Analyzers included Beckman Coulter AU5800, Siemens Advia 1800 and Roche Cobas c303 for chemistry; Beckman UniCel DXI, Siemens Atellica IM and Roche Cobas e402 for immunoassays; and two dedicated HbA1c analyzers (Bio-Rad D-100 and Tosoh G8). Using samples from the Phenome India study [9], this study provides a realistic comparison of inter-platform variability under routine laboratory conditions, offering insights critical for improving diagnostic accuracy and cross-cohort data harmonisation. Furthermore, considering the widespread prevalence of Vitamin B12 deficiency in the Indian population and the inherent challenges in its accurate diagnosis due to a lack of a universally standardised reference method, the present study placed special emphasis on Vitamin B12 measurement, a parameter known for significant method-dependent variability and inconsistent diagnostic thresholds [10–14]. Since Vitamin B12 cut-offs vary across analyzers and often fail to reflect true functional deficiency, platform-specific low Vitamin B12 thresholds were derived using functional markers such as homocysteine after adjusting for Folate. Overall, these comprehensive evaluations will support efforts towards laboratory standardisation and contribute to more accurate, reliable diagnoses and research outcomes.

## Materials and Methods

### Study Design

Phenome India: CSIR Health Cohort Knowledgebase (PI-CHeCK) cohort was implemented as a large, multi-centric field study across 37 CSIR laboratories distributed throughout India. The sampling framework targeted 10,000 participants from 17 states and 2 union territories of India, where laboratories of the Council of Scientific and Industrial Research (CSIR) are situated [9]. For this present study, 429 blood samples were randomly selected from five CSIR labs: CSIR-CFTRI Mysore (48), CSIR-CLRI Chennai (128), CSIR-IMMT Bhubaneswar (121), CSIR-NEERI Nagpur (87) and CSIR-NIScPR New Delhi (45). In total, 29 parameters were measured, of which 19 clinical chemistry analytes were tested on Beckman Coulter AU5800, Siemens Advia 1800 and Roche Cobas c303, five analytes including ApoA1, ApoB, Sodium, Potassium and Chloride were tested on Beckman Coulter AU5800 and Siemens Advia 1800 and four immunoassays on Beckman UniCel DXI, Siemens Atellica IM and Roche Cobas e402; measurement of HbA1c was performed on Bio-Rad D-100 and Tosoh G8. Samples with low volume or out-of-range values were excluded from the analysis.

### Cohort Enrolment and Sample Collection

Ethical approval was granted by the Institutional Human Ethics Committee (IHEC) of CSIR-IGIB (reference number: CSIR-IGIB/IHEC/2023-24/16) and the study was registered with the Clinical Trials Registry of India (CTRI/2024/01/061807). All participants provided written informed consent during enrolment.

### Sample Collection and processing

Blood collection followed strict standardized procedures with the National Accreditation Board for Testing and Calibration Laboratories (NABL) and the College of American Pathologists (CAP) - accredited pathology lab to ensure sample integrity. After a 10-12 hour fasting, 20 mL of blood was drawn under sterile conditions. All CSIR laboratories adhered to uniform SOPs for registration, phlebotomy, labelling, storage and transport [9]. Trained personnel and regular monitoring minimized pre-analytical variability, ensuring observed differences reflect analytical rather than sampling factors.

### Quality Control

To maintain data integrity, all analyzers were calibrated according to the manufacturer’s specifications. Daily internal quality control checks were performed using certified Bio-Rad quality control materials. Sample collection, processing and storage were conducted under controlled conditions to prevent any degradation or contamination. All analytical procedures were carried out at Healthians Laboratory (Expedient Healthcare Private Limited, a private lab accredited by NABL and CAP, at their Gurgaon, Haryana facility, India). Data entry and management were carried out using a centralised database with a double-entry verification system to minimise transcription errors. These rigorous methodological and quality assurance protocols ensured the generation of reliable, high-quality and reproducible data suitable for clinical analysis.

### Methods accuracy

The analytical accuracy of immunoassay and clinical chemistry methods on the Beckman, Siemens and Roche platforms was rigorously assessed using the BIORAD-EQAS (External Quality Assurance Scheme). Proficiency testing (PT) was conducted to verify test results through peer labs by enrolling in the Bio-Rad EQAS & CAP PT programs. Each analyzer’s results were directly compared with the Bio-Rad’s EQAS means and accuracy was evaluated using Z-score analysis and bias percentage, confirming method reliability and robust concordance with international proficiency standards.

### Statistical Analysis

Agreement among the Beckman, Siemens and Roche analyzers across 29 assays was evaluated using three complementary approaches. Passing-Bablok Regression, a non-parametric method that does not assume a specific data distribution, was used to estimate constant bias (Intercept, a) and proportional bias (Slope, b) with the best agreement indicated by A=0 and B=1 [15–17], Passing-Bablok regression was carried out using Python. Bland-Altman analysis evaluated the differences between paired measurements and their average values, to quantify the mean bias and the limits of agreement (±1.96 SD), thereby identifying any systematic deviation between methods [15, 17]. The Bland-Altman analysis was performed in Python. The Concordance Correlation Coefficient (CCC), which integrates measures of precision and accuracy, was calculated using Lin’s method in MedCalc software (Trial Version). CCC values >0.95 indicate excellent concordance, whereas values <0.90 indicate poor concordance [15, 17].

### Identifying Low Vitamin B12 Cut-off Levels

Potential cut-off values for low Vitamin B12 were identified using restricted cubic spline regression, modelling the relationship between Vitamin B12 and homocysteine, adjusted for Folate. Knot points indicated inflection points where homocysteine rose sharply, determining cut-offs for each assay. Multivariable regression analyses were conducted using Stata (version 19.5).

## Results

The method comparability study across the Beckman, Siemens and Roche platforms evaluated a comprehensive panel of clinical chemistry analytes relevant to diagnostic applications. Median values for all the assays across platforms are provided in Table 1

**Table 1:**
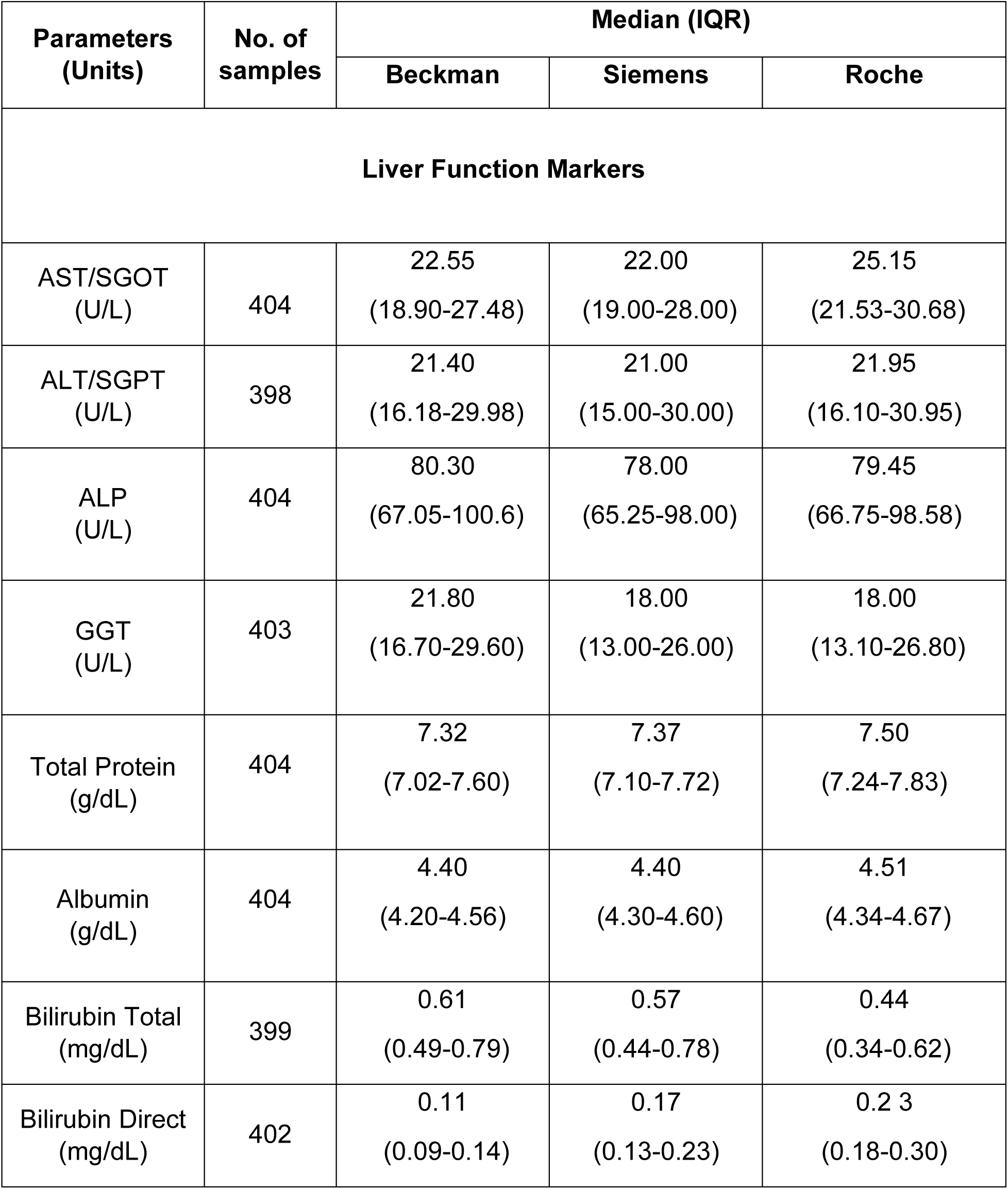
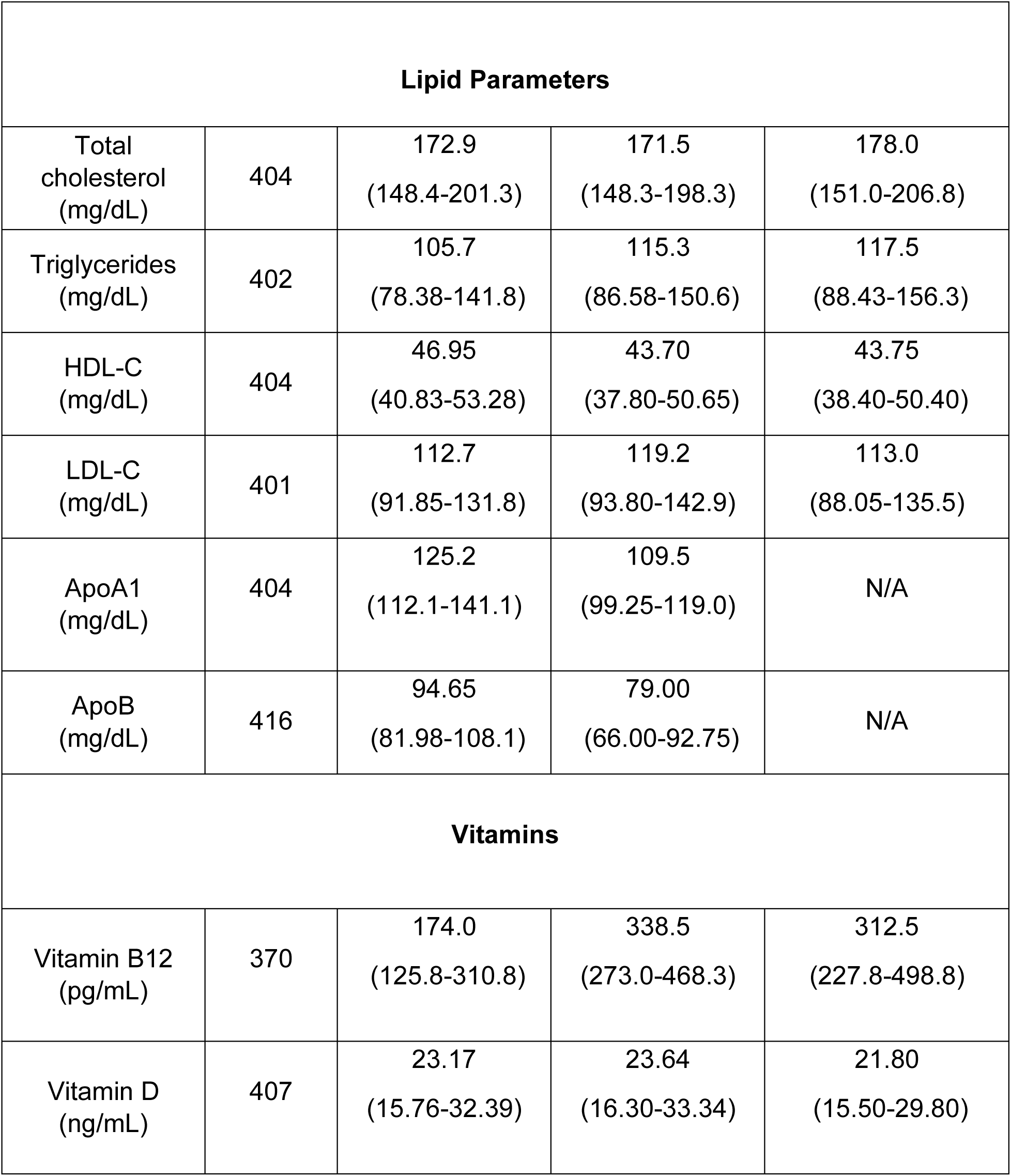
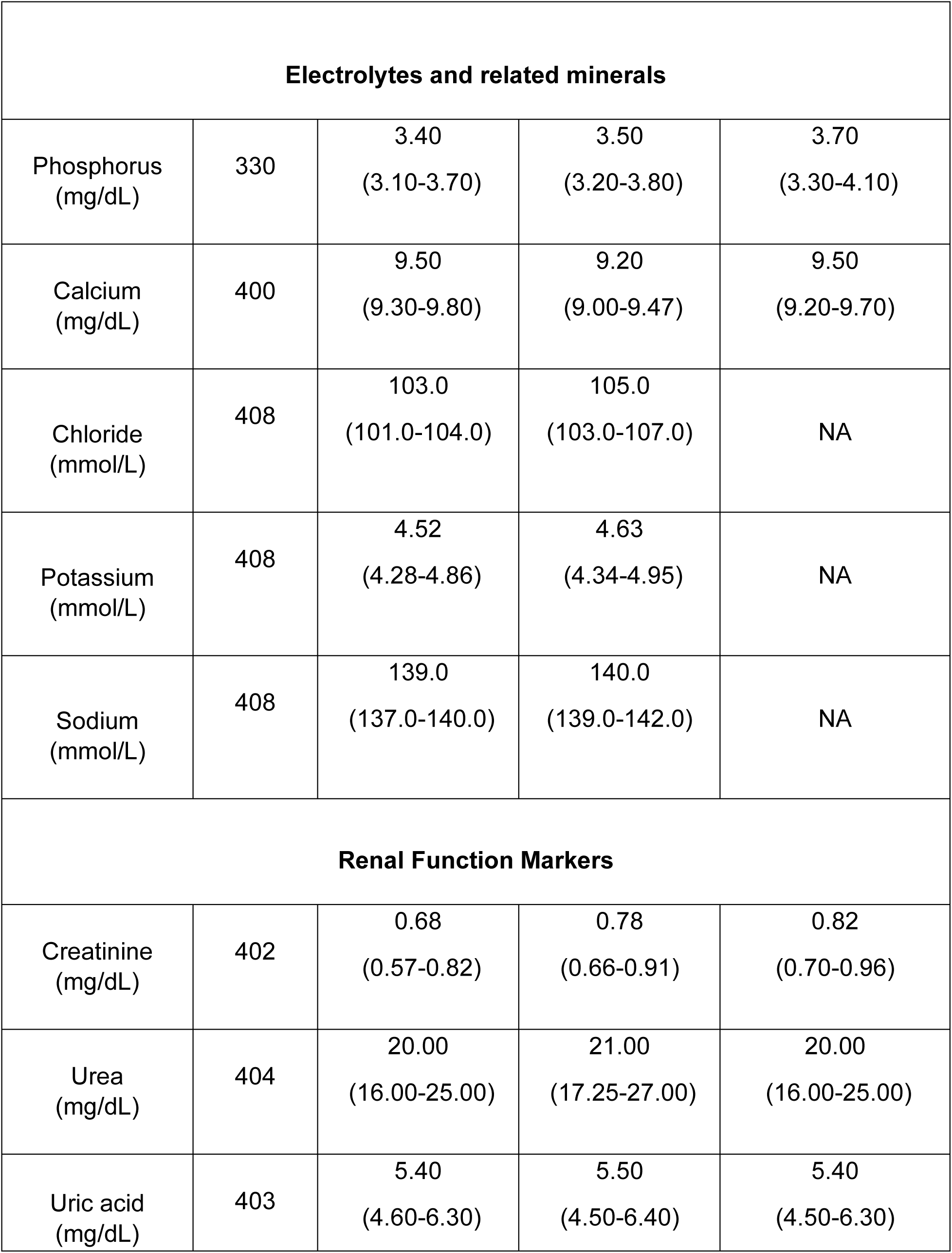
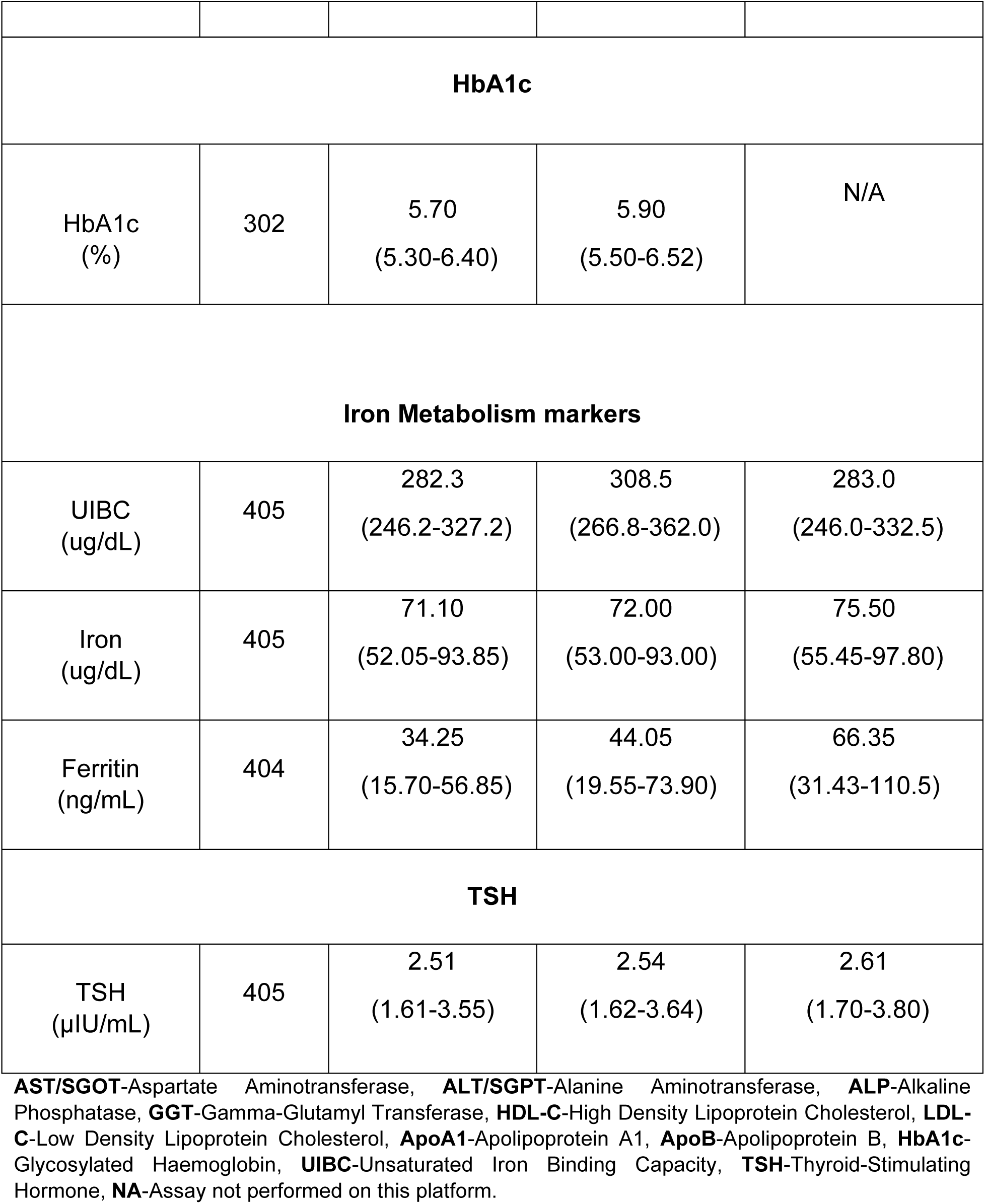
List of Clinical Chemistry & Immunoassay parameters with their Median (IQR) for the three platforms.

| Parameters<br>(Units) | No. of<br>samples | Median (IQR) |  |  |
| --- | --- | --- | --- | --- |
|  |  | Beckman | Siemens | Roche |
| Liver Function Markers |  |  |  |  |
| AST/SGOT<br>(U/L) | 404 | 22.55<br>(18.90-27.48) | 22.00<br>(19.00-28.00) | 25.15<br>(21.53-30.68) |
| ALT/SGPT<br>(U/L) | 398 | 21.40<br>(16.18-29.98) | 21.00<br>(15.00-30.00) | 21.95<br>(16.10-30.95) |
| ALP<br>(U/L) | 404 | 80.30<br>(67.05-100.6) | 78.00<br>(65.25-98.00) | 79.45<br>(66.75-98.58) |
| GGT<br>(U/L) | 403 | 21.80<br>(16.70-29.60) | 18.00<br>(13.00-26.00) | 18.00<br>(13.10-26.80) |
| Total Protein<br>(g/dL) | 404 | 7.32<br>(7.02-7.60) | 7.37<br>(7.10-7.72) | 7.50<br>(7.24-7.83) |
| Albumin<br>(g/dL) | 404 | 4.40<br>(4.20-4.56) | 4.40<br>(4.30-4.60) | 4.51<br>(4.34-4.67) |
| Bilirubin Total<br>(mg/dL) | 399 | 0.61<br>(0.49-0.79) | 0.57<br>(0.44-0.78) | 0.44<br>(0.34-0.62) |
| Bilirubin Direct<br>(mg/dL) | 402 | 0.11<br>(0.09-0.14) | 0.17<br>(0.13-0.23) | 0.2 3<br>(0.18-0.30) |

| Lipid Parameters |  |  |  |  |
| --- | --- | --- | --- | --- |
| Total cholesterol (mg/dL) | 404 | 172.9<br>(148.4-201.3) | 171.5<br>(148.3-198.3) | 178.0<br>(151.0-206.8) |
| Triglycerides (mg/dL) | 402 | 105.7<br>(78.38-141.8) | 115.3<br>(86.58-150.6) | 117.5<br>(88.43-156.3) |
| HDL-C (mg/dL) | 404 | 46.95<br>(40.83-53.28) | 43.70<br>(37.80-50.65) | 43.75<br>(38.40-50.40) |
| LDL-C (mg/dL) | 401 | 112.7<br>(91.85-131.8) | 119.2<br>(93.80-142.9) | 113.0<br>(88.05-135.5) |
| ApoA1 (mg/dL) | 404 | 125.2<br>(112.1-141.1) | 109.5<br>(99.25-119.0) | N/A |
| ApoB (mg/dL) | 416 | 94.65<br>(81.98-108.1) | 79.00<br>(66.00-92.75) | N/A |
| Vitamins |  |  |  |  |
| Vitamin B12 (pg/mL) | 370 | 174.0<br>(125.8-310.8) | 338.5<br>(273.0-468.3) | 312.5<br>(227.8-498.8) |
| Vitamin D (ng/mL) | 407 | 23.17<br>(15.76-32.39) | 23.64<br>(16.30-33.34) | 21.80<br>(15.50-29.80) |

| Electrolytes and related minerals |  |  |  |  |
| --- | --- | --- | --- | --- |
| Phosphorus<br>(mg/dL) | 330 | 3.40<br>(3.10-3.70) | 3.50<br>(3.20-3.80) | 3.70<br>(3.30-4.10) |
| Calcium<br>(mg/dL) | 400 | 9.50<br>(9.30-9.80) | 9.20<br>(9.00-9.47) | 9.50<br>(9.20-9.70) |
| Chloride<br>(mmol/L) | 408 | 103.0<br>(101.0-104.0) | 105.0<br>(103.0-107.0) | NA |
| Potassium<br>(mmol/L) | 408 | 4.52<br>(4.28-4.86) | 4.63<br>(4.34-4.95) | NA |
| Sodium<br>(mmol/L) | 408 | 139.0<br>(137.0-140.0) | 140.0<br>(139.0-142.0) | NA |
| Renal Function Markers |  |  |  |  |
| Creatinine<br>(mg/dL) | 402 | 0.68<br>(0.57-0.82) | 0.78<br>(0.66-0.91) | 0.82<br>(0.70-0.96) |
| Urea<br>(mg/dL) | 404 | 20.00<br>(16.00-25.00) | 21.00<br>(17.25-27.00) | 20.00<br>(16.00-25.00) |
| Uric acid<br>(mg/dL) | 403 | 5.40<br>(4.60-6.30) | 5.50<br>(4.50-6.40) | 5.40<br>(4.50-6.30) |
| **HbA1c** |  |  |  |  |
| HbA1c (%) | 302 | 5.70<br>(5.30-6.40) | 5.90<br>(5.50-6.52) | N/A |
| **Iron Metabolism markers** |  |  |  |  |
| UIBC (ug/dL) | 405 | 282.3<br>(246.2-327.2) | 308.5<br>(266.8-362.0) | 283.0<br>(246.0-332.5) |
| Iron (ug/dL) | 405 | 71.10<br>(52.05-93.85) | 72.00<br>(53.00-93.00) | 75.50<br>(55.45-97.80) |
| Ferritin (ng/mL) | 404 | 34.25<br>(15.70-56.85) | 44.05<br>(19.55-73.90) | 66.35<br>(31.43-110.5) |
| **TSH** |  |  |  |  |
| TSH (μIU/mL) | 405 | 2.51<br>(1.61-3.55) | 2.54<br>(1.62-3.64) | 2.61<br>(1.70-3.80) |

**AST/SGOT**-Aspartate Aminotransferase, **ALT/SGPT**-Alanine Aminotransferase, **ALP**-Alkaline Phosphatase, **GGT**-Gamma-Glutamyl Transferase, **HDL-C**-High Density Lipoprotein Cholesterol, **LDL-C**-Low Density Lipoprotein Cholesterol, **ApoA1**-Apolipoprotein A1, **ApoB**-Apolipoprotein B, **HbA1c**-Glycosylated Haemoglobin, **UIBC**-Unsaturated Iron Binding Capacity, **TSH**-Thyroid-Stimulating Hormone, **NA**-Assay not performed on this platform.

Passing-Bablok regression slopes detailing proportional bias and inter-assay agreement are visualized as forest plots for 29 parameters (Figure 1 and Supplementary Table S1). Concordance correlation coefficients (CCC) and pairwise analyzers comparisons are visualized as forest plots in Figure 2 and reported in Supplementary Table S2. The CCC analysis included 29 assays across four key pairings: Beckman versus Siemens, Siemens versus Roche, Beckman versus Roche and Bio-Rad versus Tosoh for HbA1c. High concordance (CCC >0.90) was observed for major analytes such as ALT, AST, Total cholesterol and HbA1c, whereas apolipoprotein A1, ApoB, Sodium and Chloride and Vitamin B12 displayed lower concordance, indicating high platform-dependent variability. Bland-Altman plots for most parameters including ALT, AST, Total cholesterol, Urea, Uric acid and HbA1c, exhibited narrow limits and minimal systematic bias, supporting robust cross-platform interchangeability (Supplementary Figure S1-S7, Supplementary Table S3). In contrast, broader limits and greater proportional differences were noted for UIBC, Ferritin, Vitamin B12, and apolipoproteins, including ApoA1 and ApoB, reflecting greater measurement variability as shown in Figure 3. These findings confirm some degree of analyzer interchangeability for analytes with overlapping reference ranges, supporting robust clinical decision making across platforms.

**Figure 1.**
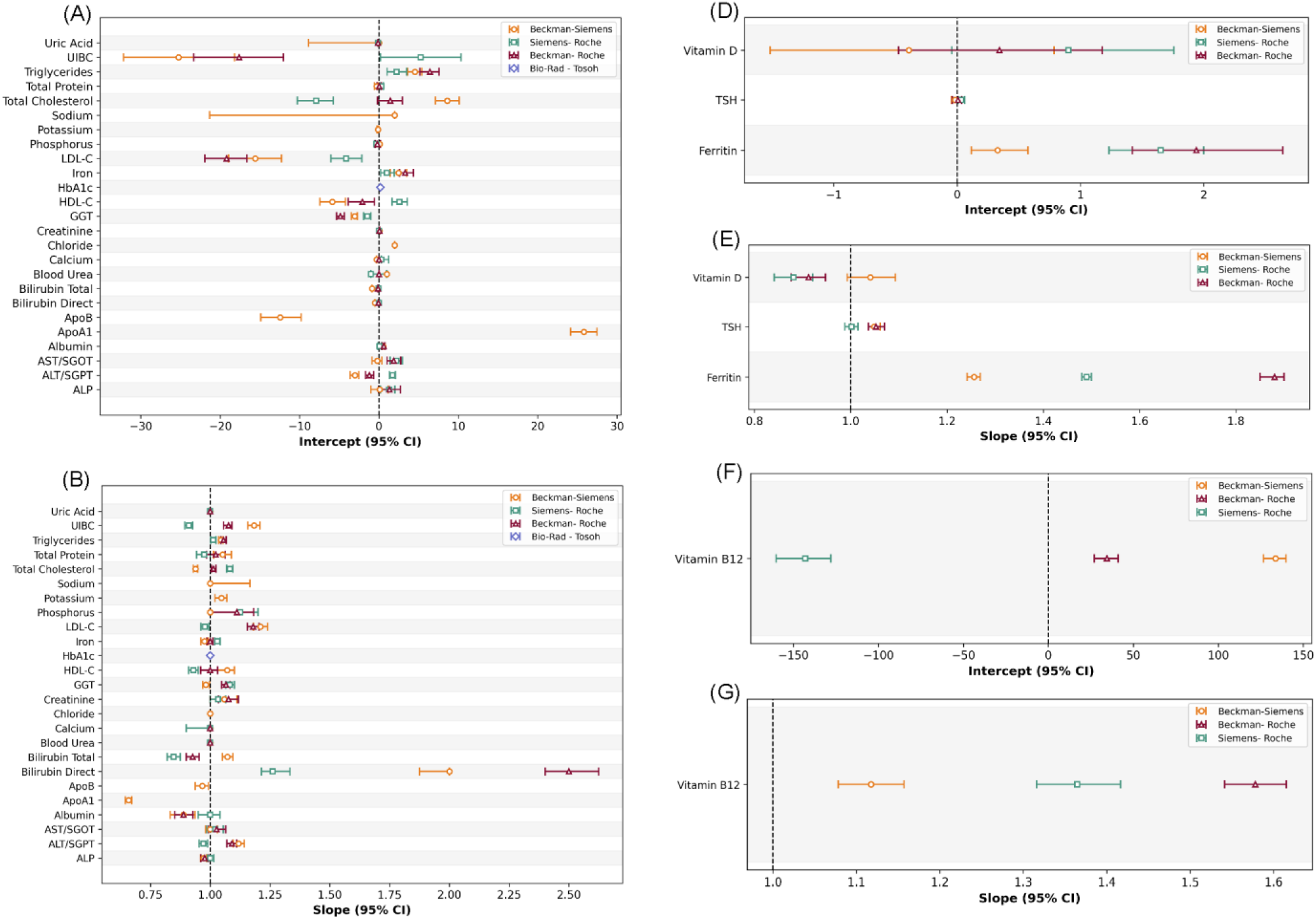
Passing-Bablok regression analysis of inter-analyzer performance. Figure A and B display intercept and slope estimates (with 95% CI) for 25 routine clinical chemistry assays across three analyzer pairs: Beckman-Roche (maroon), Siemens-Roche (green) Beckman-Siemens (orange); Bio-Rad-Tosoh (slate blue) is shown for HbA1c. Each point represents one assay’s regression parameter, highlighting systematic (intercept) and proportional (slope) biases. Figure C and D show corresponding estimates for three immunoassays, while Figure E and F focus on Vitamin B12. Dashed lines indicate identity (intercept = 0, slope = 1) for reference. **\*Comparison for ApoA1, ApoB, Sodium, Potassium, Chloride and HbA1c was performed between two analyzers.**

**Figure 2.**
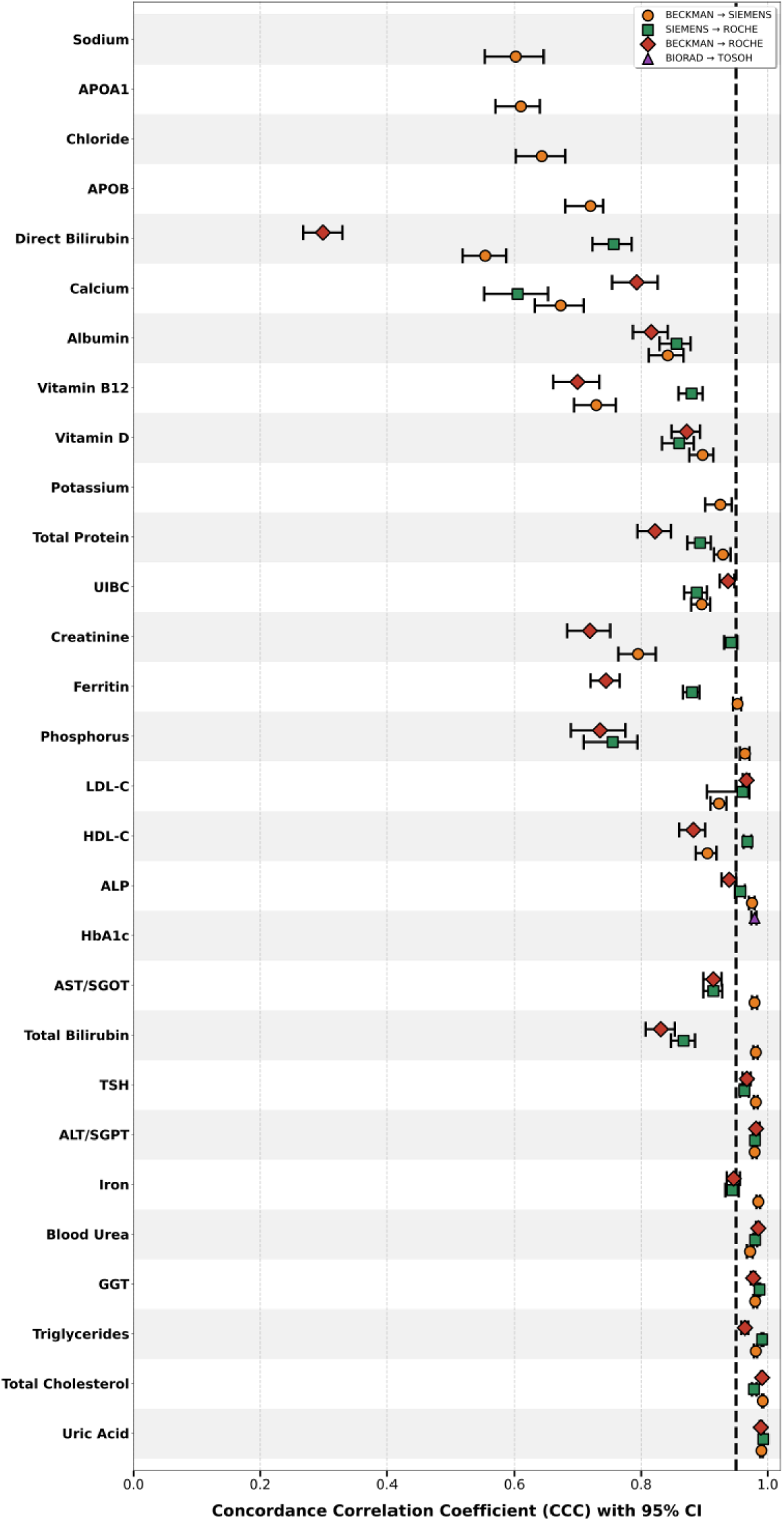
Comparison of Concordance Correlation Coefficients (CCC) Across Analyzers. This figure displays the CCC for 25 clinical chemistry and four immunoassay parameters measured across three analyzer pairings: Beckman-Roche (maroon), Siemens-Roche (green), Beckman-Siemens (orange) and HbA1c was performed between Bio-Rad and Tosoh (slate blue). Each point represents the CCC value for a specific analyte, indicating the degree of agreement between instrument platforms. Analytes are ordered from highest to lowest concordance across the x-axis. Higher CCC values (closer to 1) reflect stronger agreement between methods, with most analytes showing CCC ≥ 0.85 across pairs, while a subset including Sodium, Chloride, Direct Bilirubin, ApoB and ApoA1 exhibits comparatively lower agreement. The dotted black line denotes a CCC value of 0.95, indicating the predefined threshold for acceptable concordance. **\*Comparison for ApoA1, ApoB, Sodium, Potassium, Chloride and HbA1c was performed between two analyzers**.

**Figure 3.**
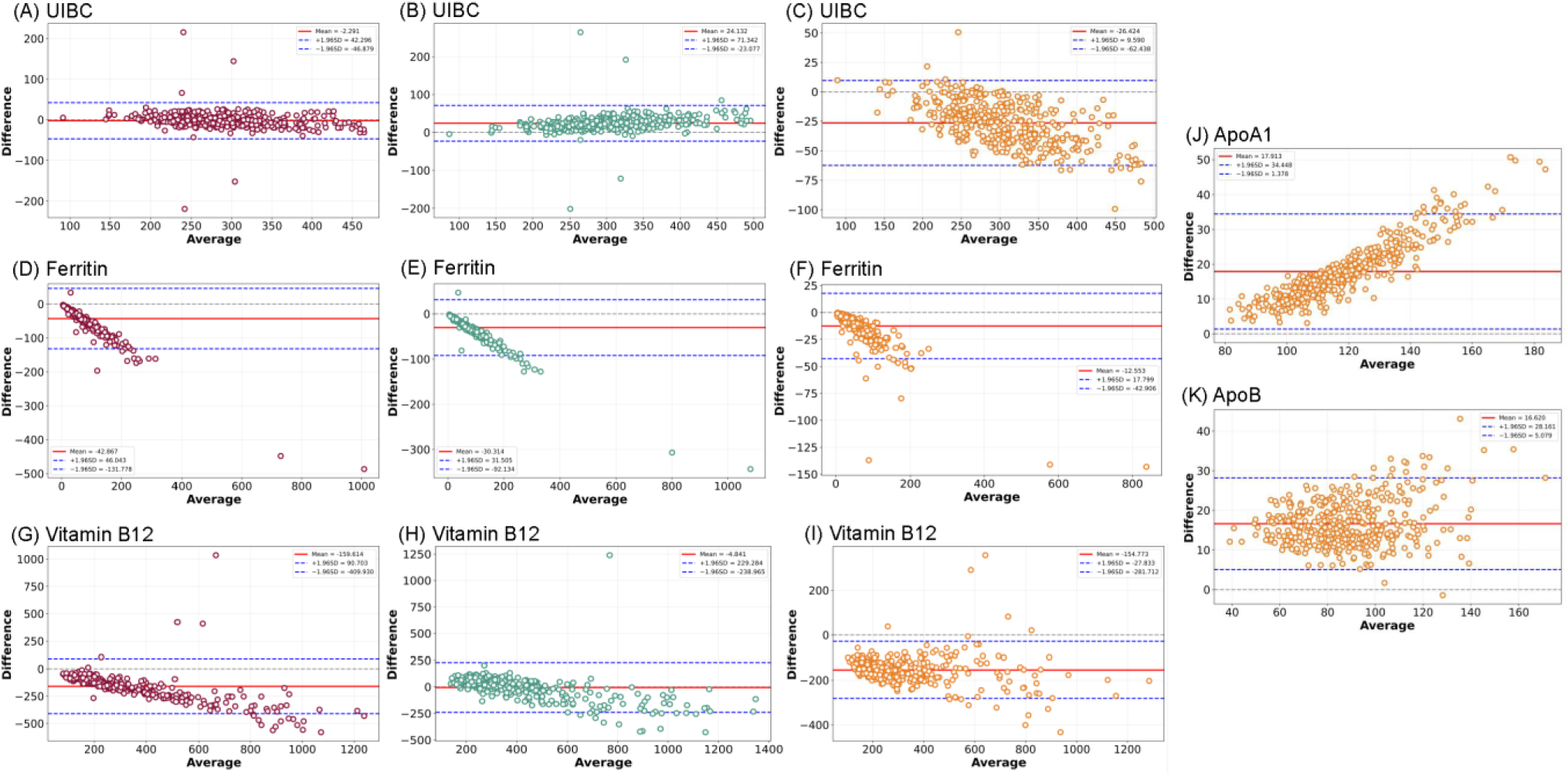
Bland-Altman analysis of agreement between clinical chemistry and immunoassay analyzers. Bland-Altman plots displaying the mean bias and 95% limits of agreement for analytes measured across the compared analyzers. Each figure shows comparisons for different parameters: UIBC, Ferritin, Vitamin B12, ApoA1 and ApoB. Horizontal lines denote the mean difference (bias) as well as upper and lower limits of agreement (mean±1.96 SD). Three analyzer pairings: Beckman-Roche (maroon), Siemens-Roche (green), Beckman-Siemens (orange). *Average values were calculated as the mean of paired methods: 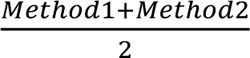. Mean differences were determined using the formula: where (*Method*1 − *Method*2) correspond to: Beckman-Roche (Beckman as Method1, Roche as Method2), Beckman-Siemens (Beckman as Method1, Siemens as Method2), Siemens-Roche (Siemens as Method1, Roche as Method2).

### Liver Function Markers

Passing-Bablok regression analysis of enzymatic markers including ALT, AST, ALP and GGT showed a systematic bias across platforms, whereas Direct Bilirubin showed a proportional bias as shown in Figure 1. The concordance correlation coefficient analysis demonstrated good to excellent agreement for ALT, AST, ALP and GGT. Total Bilirubin demonstrated overall poor agreement, except for the Beckman-Siemens comparison, which showed excellent agreement. In contrast, Direct Bilirubin and Albumin exhibited marked inter-platform variability with poor CCC values (Figure 2). Bland-Altman analyses showed minimal mean difference between these platforms, indicating no clinically relevant systematic bias and good agreement across the measurement range, as shown in Supplementary Table S3.

### Electrolytes and related minerals

Calcium showed moderate inter-platform agreement with Passing-Bablok slopes close to unity and relatively narrow Bland-Altman limits, yet the CCC was 0.673, indicating poor concordance. In contrast, Potassium demonstrated good concordance (CCC 0.925; intercept −0.11, slope 1.04), whereas Sodium and Chloride, despite intercepts of 2 and slopes of 1, had poor CCC values (0.602 and 0.643), indicating poor agreement. Overall, electrolytes and related minerals exhibited some of the lowest concordance values among analyte groups, reflecting notable inter-platform discrepancies (Figure 2) and Supplementary Tables S1, S2 and S3.

### Renal Function Markers

Serum Creatinine showed inter-platform agreement, with slopes near unity and minimal intercept bias as shown in Figure 1. Urea and Uric acid demonstrated excellent agreement with slopes close to unity and high concordance (Figure 2). CCC analysis of these parameters revealed good to excellent agreement for Urea and Uric acid (CCC 0.97-0.99), except for Creatinine, which showed moderate to poor concordance (CCC 0.71-0.94) as represented in Figure 2. Bland-Altman analysis demonstrated that the mean difference between these platforms is close to zero except for Urea, which showed a mean difference of −1.57 for Beckman-Siemens and 1.24 for Siemens-Roche comparisons, indicating no clinically relevant systematic bias.

### Lipid Parameters

Passing-Bablok regression analysis revealed a positive bias in the HDL-C measurements in the Siemens-Roche comparison. For Total cholesterol, a positive constant bias was observed in both the Beckman-Siemens and Beckman-Roche comparisons. The slope was unity for Triglycerides across all analytical platforms, indicating no proportional bias. In contrast, for HDL-C and Total cholesterol, a unit slope was observed only in the Beckman-Siemens and Siemens-Roche comparisons. ApoA1 demonstrated a marked positive systematic (constant) bias accompanied by proportional bias, whereas ApoB showed a systematic negative bias in the Beckman-Siemens comparison. For LDL-C, all three analyzers exhibited a systematic negative bias, with slopes close to unity, indicating minimal proportional bias (Figure 1). Concordance correlation analyses for both LDL-C and HDL-C showed moderate to good agreement, whereas agreement was poor for the apolipoproteins Figure 2. Bland Altman analysis for LDL-C and HDL-C showed no proportional bias; however, for ApoA1 and ApoB comparisons, most observations clustered around a distinct mean difference, indicating that Beckman consistently yielded higher values than Siemens across the measurement range, with wide limits of agreement reflecting considerable variability, as shown in Figure 3(J) ApoA1 and 3 (K) ApoB.

### Iron Metabolism Markers

Among markers of Iron metabolism, Passing-Bablok regression analyses for Iron and UIBC showed a systematic bias across analyzers. For Ferritin, the Beckman-Siemens comparison showed a slight proportional bias (slope 1.2), whereas other comparison pairs showed both systematic and proportional bias (Figure 1). Concordance correlation analyses for Ferritin showed good agreement between Beckman and Siemens, whereas other inter-platform comparisons exhibited poor agreement. Iron displayed overall good agreement (CCC 0.94-0.98). In contrast, UIBC measurements displayed poor agreement except for the Beckman-Roche comparison, which showed moderate agreement (Figure 2). The Bland-Altman analysis further supported these findings as shown in Figure 3(A), 3(B) and 3(C) for UIBC; 3(D), 3(E) and 3(F) for Ferritin.

### Vitamins

Combined method-comparison analysis revealed substantial inter-analyzer disagreement for Vitamin B12, with Passing-Bablok regression indicating significant systematic and proportional bias across all pairs of analyzers (Figure 1). These findings were supported by a Bland-Altman analysis, which showed large mean biases and wide limits of agreement as shown in Figure 3(G), 3(H) and 3(I) Vitamin B12, as well as concordance correlation coefficients indicating a poor agreement (CCC: 0.699-0.880, Figure 2). In contrast, Vitamin D demonstrated moderate analytical agreement across platforms, with minimal constant bias, narrower limits of agreement, and consistently poor concordance (CCC: 0.860–0.897) (Figure 1 and 2)

### HbA1c

HbA1c measurements showed near-perfect agreement between Bio-Rad and Tosoh analyzers, with Passing-Bablok slopes of 1.0 and negligible intercept bias (Figure 1). Concordance for HbA1c was perfect (CCC ∼1.0), with minimal difference and narrow Bland-Altman limits, supporting robust interchangeability (Figure 2 and Supplementary Table S3).

### TSH

TSH assays demonstrated strong inter-platform agreement, with slopes near unity and minimal intercept bias, as shown in Figure 1. High Concordance coefficients exceeded 0.95, along with narrow Bland-Altman limits of agreement, indicate clinically acceptable interchangeability among the tested platforms (Figure 2 and Supplementary Table S3).

### Thresholds defining low B12 status

Restricted cubic spline regression analyses revealed a non-linear pattern between Vitamin B12 and Homocysteine, with each platform showing a distinct inflection point at which Homocysteine levels rose sharply as Vitamin B12 declined. These inflection points represent physiological thresholds below which Vitamin B12 appears insufficient. The estimated cut-offs were ∼100 pg/mL for Beckman Coulter, ∼280 pg/mL for Siemens and ∼330 pg/mL for Roche. Prediction curves derived from the spline fits, along with their corresponding 95% confidence intervals, supported the robustness of these inflection points (Figure 4). These findings highlight platform-dependent variability in the Vitamin B12-homocysteine relationship and underscore the need for assay-specific reference thresholds.

**Figure 4.**
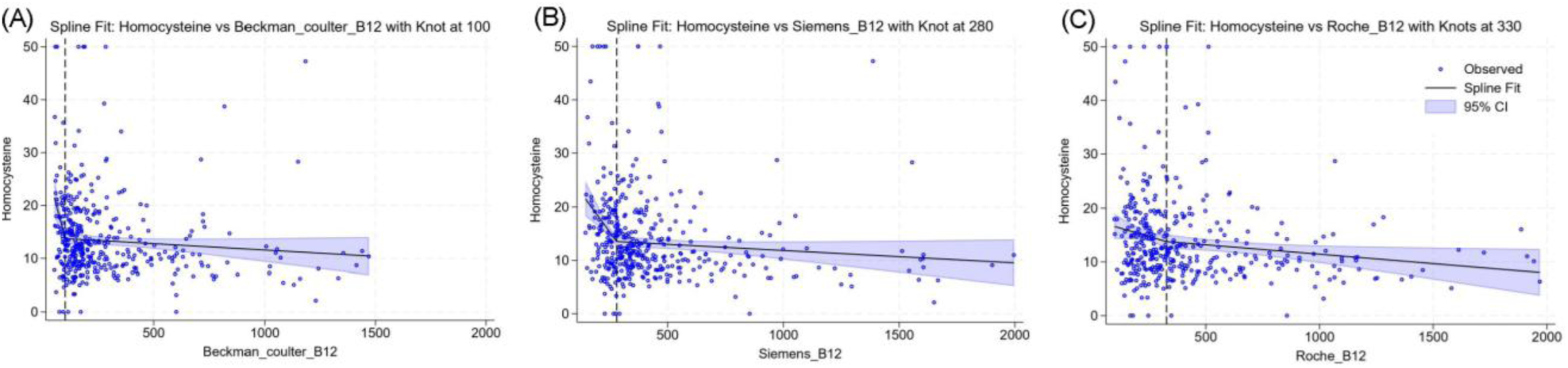
Multivariable regression analysis of the association between serum homocysteine and Vitamin B12 across analytical platforms. Scatterplots with fitted multivariable regression lines illustrate the relationship between Serum Homocysteine (Y-axis, µmol/L) and Vitamin B12 concentrations (X-axis, pg/mL) after adjustment for Folate levels. Analyses were conducted separately using platform-specific Vitamin B12 measurements for: (A) Beckman Coulter, (B) Siemens and (C) Roche. Cut-off value of 100 pg/mL was observed for Beckman, 280 pg/mL for Siemens and 330 pg/mL for Roche analyses.

## Discussion

This study significantly expands on earlier investigations, which often focused on a limited set of analytes or examined only one analytical category. By evaluating both clinical chemistry and immunoassay parameters across three major automated platforms-Beckman, Siemens and Roche, this work provides a comprehensive and integrated assessment of Inter-platform comparability. As correlation analysis alone does not adequately assess agreement or identify bias, this study applied a robust multi-metric approach, including Passing-Bablok regression, Bland-Altman plots and CCC, to assess both systematic and proportional biases. This approach allows for simultaneous evaluation of variability across multiple assay classes rather than studying them in isolation, thus offering a more holistic understanding of inter-platform performance. Our findings showed considerable variability in inter-platform agreement across assay classes. Among enzymatic markers, ALT, AST and ALP demonstrated high concordance consistent with previous reports showing that enzymatic assays for ALT, AST and ALP are generally robust and interchangeable across popular platforms [6, 18]. A previous hospital-based study reported high variability in Chloride measurements between Nova Stat and Vitro XT analyzers, while Sodium and Potassium showed good consistency [19]. Conversely, another study found good agreement for Chloride using Beckman Coulter AU480 and ExDs analyzers [20]. These discrepancies highlight the need for caution, as inter-platform variability may affect clinical decisions. Renal function markers showed variable inter-platform agreement, with Creatinine showing moderate agreement across analyzers. Such discrepancies, although modest, may affect the interpretation of eGFR-based results in diseases such as chronic kidney disease [21]. On the other hand, Urea and Uric acid showed excellent agreement across all analyzers (CCC > 0.98). Bikila et al. reported a positive bias in Urea measurements between the Dirui DR-7000 and the Roche Cobas 6000, attributed to differences in wavelength and reagents [22]. Mishra et al. found a strong correlation but significant differences in Uric acid measurements between Selectra and Vitros 350 [23]. A hospital-based study reported high agreement for renal parameters, including Urea, Uric acid and Creatinine, using ERBA Chem 7 and Selectra Pro M [24]. These findings highlight the importance of calibration and traceability in ensuring renal function comparability across different platforms. Lipid profile parameters exhibited substantial inter-platform variability. LDL-C showed systematic bias, which could affect cardiovascular risk assessment. Total cholesterol and Triglycerides correlated strongly between platforms, while HDL-C agreement was slightly less but still acceptable. Similar patterns have been reported in prior population-based and cross-analyzer studies [25–27]. Apolipoproteins (ApoA1 and ApoB) showed the poorest comparability (CCC 0.40-0.72). Similar findings were reported for ApoB in the Multi-Ethnic Study of Atherosclerosis (MESA) study, which highlighted inter-assay bias due to non-harmonised reagents [28]. In contrast, other comparative studies of ApoA1 and ApoB using different analyzers revealed good correlation between measurement methods [29, 30]. Serum protein assays showed systematic biases: Beckman underestimates Albumin and slightly overestimates Total Protein, whereas Siemens and Roche are more consistent. These differences are likely method-dependent, since Albumin assays vary by dye-binding method (bromocresol green vs. bromocresol purple) or by immunoturbidimetry, leading to manufacturer-specific bias [31]. Bilirubin, particularly Direct Bilirubin, exhibited marked inter-platform variability. Previous studies have also reported significant inter-analyzer differences, including a positive bias on Vitros compared to AU680 [32] and discrepancies between the Roche Cobas 6000 and Olympus AU2700, attributed to delta bilirubin in patients with Hyperbilirubinemia [33]. Serum Iron showed moderate to good inter-platform agreement. Previously, a comparison of Iron measurements using Dimension RxL and Vitros 950 analyzers suggested good comparability and interchangeability across platforms [34]. Differences in UIBC were more pronounced. These variations likely arise from distinct assay methodologies and reagent formulations and indicate caution when comparing Iron metabolism parameters across platforms. Among the immunoassay parameters, Ferritin and Vitamin B12 showed the greatest inter-platform disagreement. Although Ferritin measurements showed strong concordance between the Beckman and Siemens platforms (CCC of −0.95), there was a substantial overestimation on the Roche platform, which may reflect differences in antibody specificity or calibrator traceability. Similar discrepancies have been documented previously when comparing Ferritin assays on the Abbott, Roche and Vitros analyzers [35, 36]. These observations highlight that high correlation does not necessarily imply analytical interchangeability, as both proportional and constant biases can significantly influence clinical interpretation. Vitamin B12 demonstrated notably poor commutability, characterised by wide inter-analyzer variability and low CCC values. The highest correlation was observed between Roche and Siemens platforms (CCC of 0.87). A multi-centre evaluation reported strong inter-assay correlation but identified the poorest agreement between Beckman and Siemens [13]. This aligns with our findings, which showed poor agreement between Beckman and Siemens analyzers for Vitamin B12 measurement. Such inconsistencies highlight the analytical vulnerability of Vitamin B12 assays and the potential implications for clinical interpretation, particularly in defining deficiency thresholds. In the present study, platform-specific inflection points for low Vitamin B12 status were obtained using homocysteine as a functional marker. The thresholds varied substantially across platforms, reinforcing the lack of a universally applicable cut-off. This variability has significant clinical consequences, as reliance on absolute Vitamin B12 concentrations without accounting for assay-specific bias may lead to misclassification of deficiency in both clinical and epidemiological contexts. Consistent with our results, a population-based study found that Vitamin B12 reference ranges were similar between Roche and Siemens, whereas Beckman showed significantly lower values, with its intervals not aligning with the manufacturer’s references [37]. To minimise these risks, laboratories should consider reporting not only the measured Vitamin B12 concentration but also the analytical platform and manufacturer details. Such transparency would enhance interpretative accuracy and promote more consistent patient management across different testing environments. Vitamin D assays (25-Hydroxy Vitamin D) showed moderate agreement (CCC 0.86=0.90) with Roche consistently reporting lower values than Siemens and Beckman, resembling prior reports attributing differences to antibody specificity, incomplete isoform cross-reactivity and calibration issues leading to misclassification of deficiency. [38, 39]. TSH and HbA1c assays demonstrated excellent inter-platform agreement, with near-unity slopes and narrow variation. These results align with previous studies across different immunoassay platforms, confirming their wide reliability and interchangeability. [40, 41].

Key strengths of this study include extensive assay coverage across multiple analyzers, particularly immunoassays, and robust multivariate statistical analyses. One limitation of this study was the absence of complementary functional Vitamin B12 markers, like Methylmalonic acid, for more precise deficiency thresholds.

## Conclusion

We advocate a standard policy requiring laboratories to report the instrument used for testing, a practice already adopted by only a few centres. Enzymatic markers, Urea, Uric acid, and TSH demonstrated strong inter-analyzer agreement, while several important clinical assays, including electrolytes, direct Bilirubin, Ferritin, UIBC, Vitamin B12 and apolipoproteins, showed poor concordance. Lipid measurements and Iron-binding assays also revealed substantial variability. These differences highlight the need for analyzer-specific reference intervals, cautious interpretation of results in longitudinal patient monitoring and the urgent need for harmonisation initiatives, particularly for Vitamin assays. Our cut-off analysis further underscores that platform dependency profoundly influences clinical classification of Vitamin B12 deficiency, with major implications for both patient care and population-based studies.

## Data Availability

Anonymized data for public use may be made available after 3 years from completion of the baseline phase of the study or as per advisory from the Monitoring Committee of the project if any revisions are required.

## Acknowledgments

We acknowledge the support from CSIR, participants and volunteers. Further, the following names are acknowledged: Rakesh Kanakrajan Vijyakumari (**CSIR-CCMB**), Kavya Manjula Gurubasavaiah (**CSIR-CFTRI**), Vandhana Anumaiya (**CSIR-CLRI**), Rohit Kumar **(CSIR-IGIB**), Chandrama Dipa, Adyasha Bijay Mishra (**CSIR-IMMT**), Bhagyashree Likhitkar, Preshita Bhatt (**CSIR-NCL**), Sneha Joshi (**CSIR-NEERI**), Sohini Sengupta **(Healthians Labs).**

## Author Contributions

Phenome India Consortium Authors

## Study Planning and Conceptualization

Viren Sardana, Shantanu Sengupta, Kumardeep Chaudhary (**CSIR-IGIB**)

## Project conceptualization

Shantanu Sengupta, Debasis Dash, Viren Sardana, Kumardeep Chaudhary, Ajay Pratap Singh (**CSIR-IGIB**), Giriraj Ratan Chandak, Swasti Raychaudhuri, Karthik Bharadwaj Tallapaka (**CSIR-CCMB**), Mahesh J Kulkarni, Mahesh Dharne (**CSIR-NCL**), Partha Chakraborty, Dipyaman Ganguly, (**CSIR-IICB**), Umakanta Subudhi (**CSIR-IMMT**).

## Collection management

Shantanu Sengupta, Debasis Dash, Viren Sardana, Kumardeep Chaudhary, Vamsi K. Yenamandra, Aastha Mishra, Ajay Pratap Singh, Swarnendu Bag, Pankaj Pandey, Ankita Sahu, Komal Jindal, Vivek Junghare, Tarani Mathur, Meghana Arvind, Satyartha Prakash, Vignesh S Kumar, Deepak (**CSIR-IGIB**), Swasti Raychaudhuri, Giriraj Ratan Chandak, Karthik Bharadwaj Tallapaka, Rakhesh K V (**CSIR-CCMB**), Ashok P Giri, Narendra Y Kadoo, Mahesh J Kulkarni, Dhanasekaran Shanmugam, Mahesh S Dharne, Syed G Dastager, Chiranjit Chowdhury, Vamkudoth KR (Koteswara Rao, has actively contributed during sample collection) (**CSIR-NCL**), Partha Chakraborty, Dipyaman Ganguly, Shilpak Chatterjee (**CSIR-IICB**), Umakanta Subudhi, Bhabani S Jena, Trupti Das, Boopathy Ramasamy, T Pavan Kumar (**CSIR-IMMT**), Prakash M Halami, S P Muthukumar (**CSIR-CFTRI**), Suresh Kumar Anandasadagopan (**CSIR-CLRI**), Shilpa Paranjape, Prashanti Niwant (**CSIR-NEERI**), Romi Wahengbam, Tridip Phukan, Pankaj Bharali, Prasenjit Manna (**CSIR-NEIST**), Arvind Meena, Arun Uniyal (**CSIR-NIScPR**),

## Portal development and management

Kumardeep Chaudhary, Debasis Dash, Viren Sardana, Ajay Pratap Singh, Shantanu Sengupta, Satyartha Prakash, Vignesh S Kumar, Anshul Verma, Safeer Khan, Anshul Bhardwaj (**CSIR-IGIB**), Gopal Krishna Patra (**CSIR-4PI**), Nikhilesh Yadav (**CSIR-NCL**), Abbani Rakesh (**CSIR-NAL**).

## Sample Collection/Assays

Ankita Sahu, Shilpa Ray, Bharti Sharma, Mohammad Azhar Uddin, Rohit Kumar, Swati Bagdi, Sumant kumar, Abhishek Kumar, Shubham Kumar (**CSIR-IGIB**), MK Kanakavalli, Rakesh Kanakrajan Vijyakumari, Neha Kumari, Dibya Rana Saha Roy (**CSIR-CCMB**), Kavya Manjula Gurubasavaiah **(CSIR-CFTRI)**, Milind Sanjay Kale, Bhagyashree Likhitkar, Preshita Bhatt (**CSIR-NCL**), Saheli Chowdhury, Pratitusti Basu (**CSIR-IICB**), Adyasha Bijay Mishra, (**CSIR-IMMT**), Vandhana Anumaiya (**CSIR-CLRI**), Manuj Kr Das(**CSIR-NEIST**), Akshika (**CSIR-NIScPR**), Sneha Joshi **(CSIR-NEERI).**

## Data management and curation

Viren Sardana, Shantanu Sengupta, Kumardeep Chaudhary, Md. Intyaz Ali, Mamta Rathore, Satyartha Prakash (**CSIR-IGIB**)

## Manuscript writing, review and editing

Viren Sardana, Shantanu Sengupta, Rajat Ujjainiya, Mamta Rathore, Md. Intyaz Ali (**CSIR-IGIB**)

## Statistical Analysis and Data Visualization

Viren Sardana, Shantanu Sengupta, Kumardeep Chaudhary, Md. Intyaz Ali, Satyartha Prakash, Mamta Rathore (**CSIR-IGIB**), Ashish Awasthi (**CSIR-CDRI**)

## Supplementary Material

### Supplementary Tables

**Table S1:**
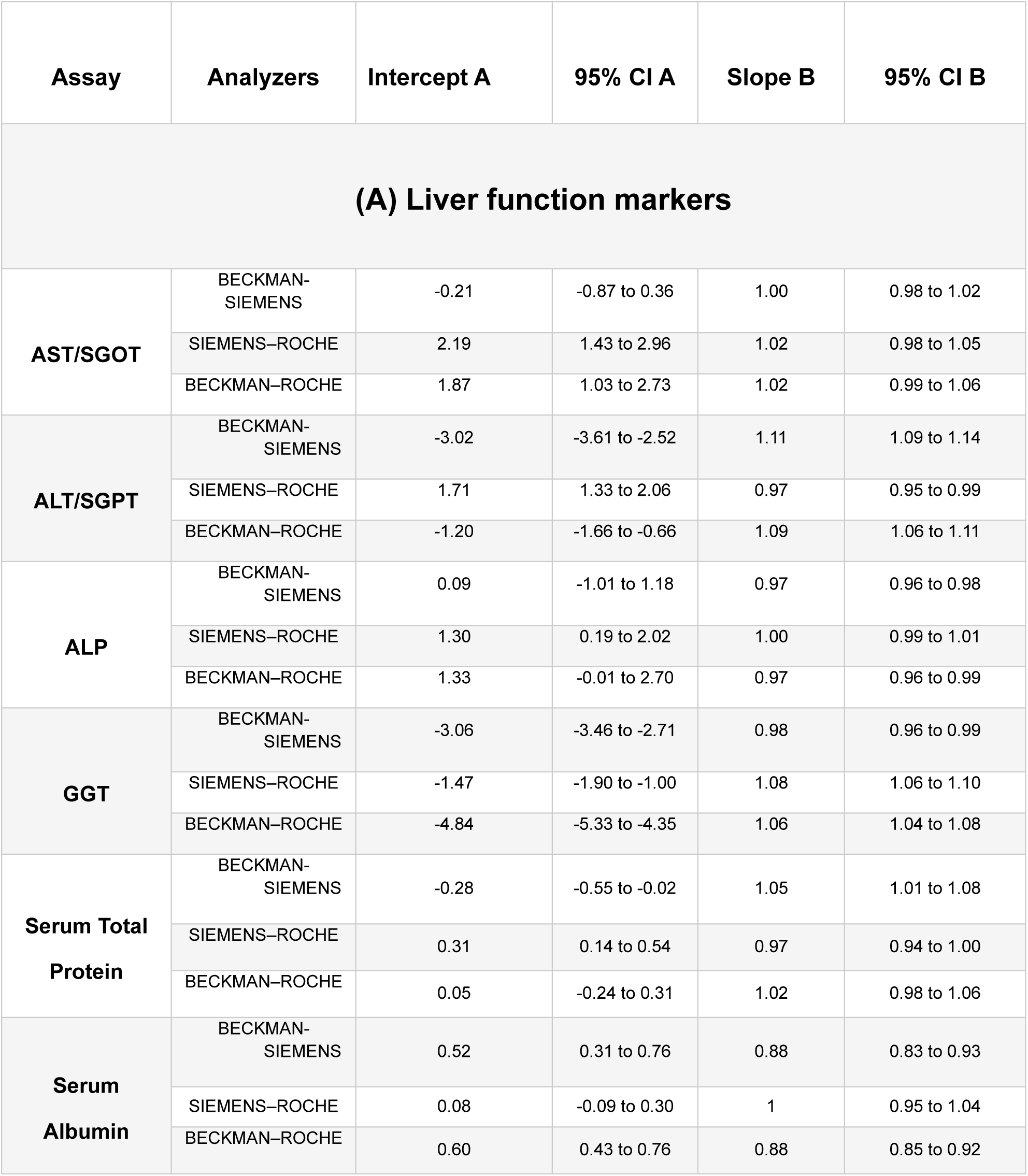
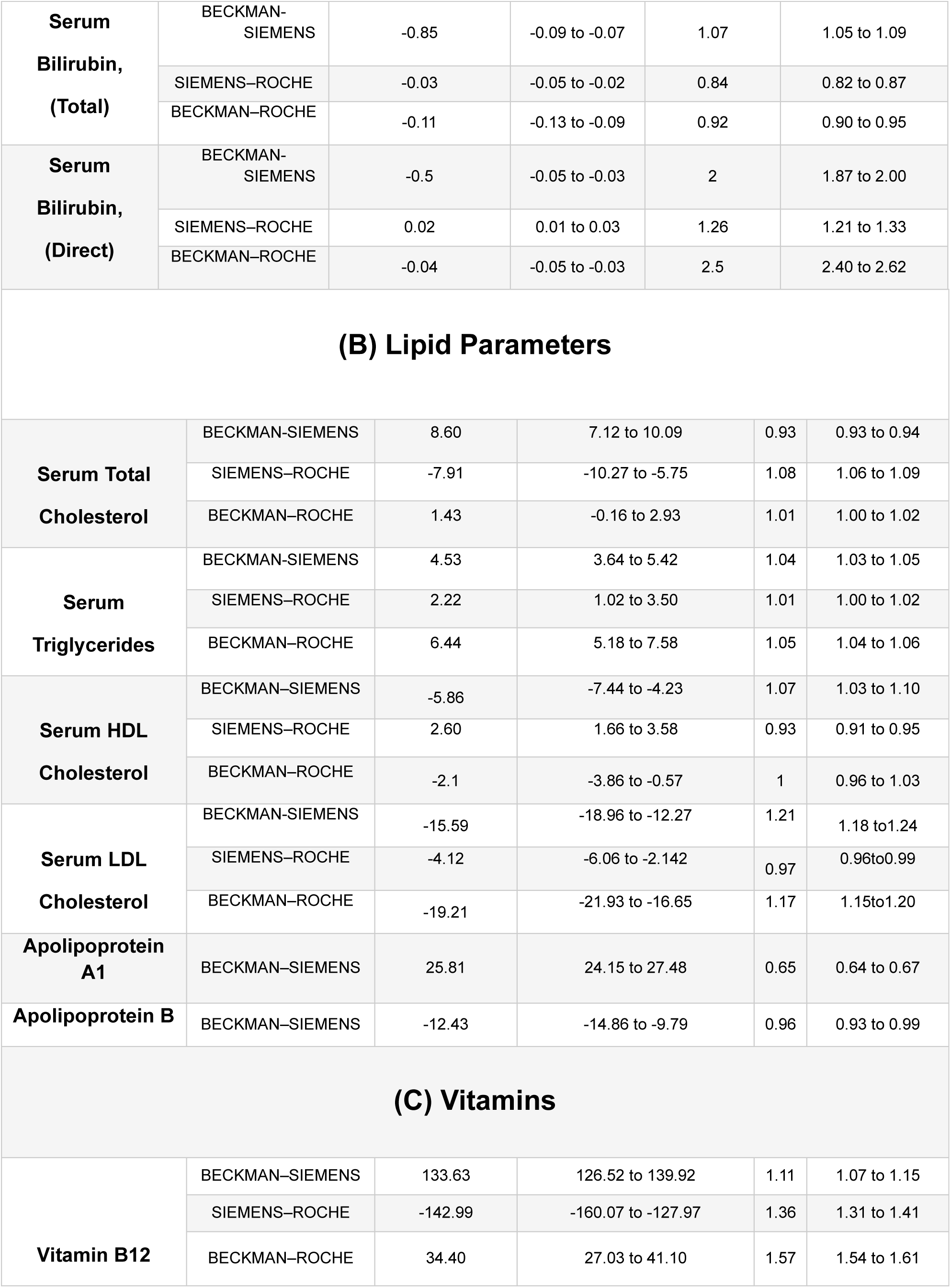
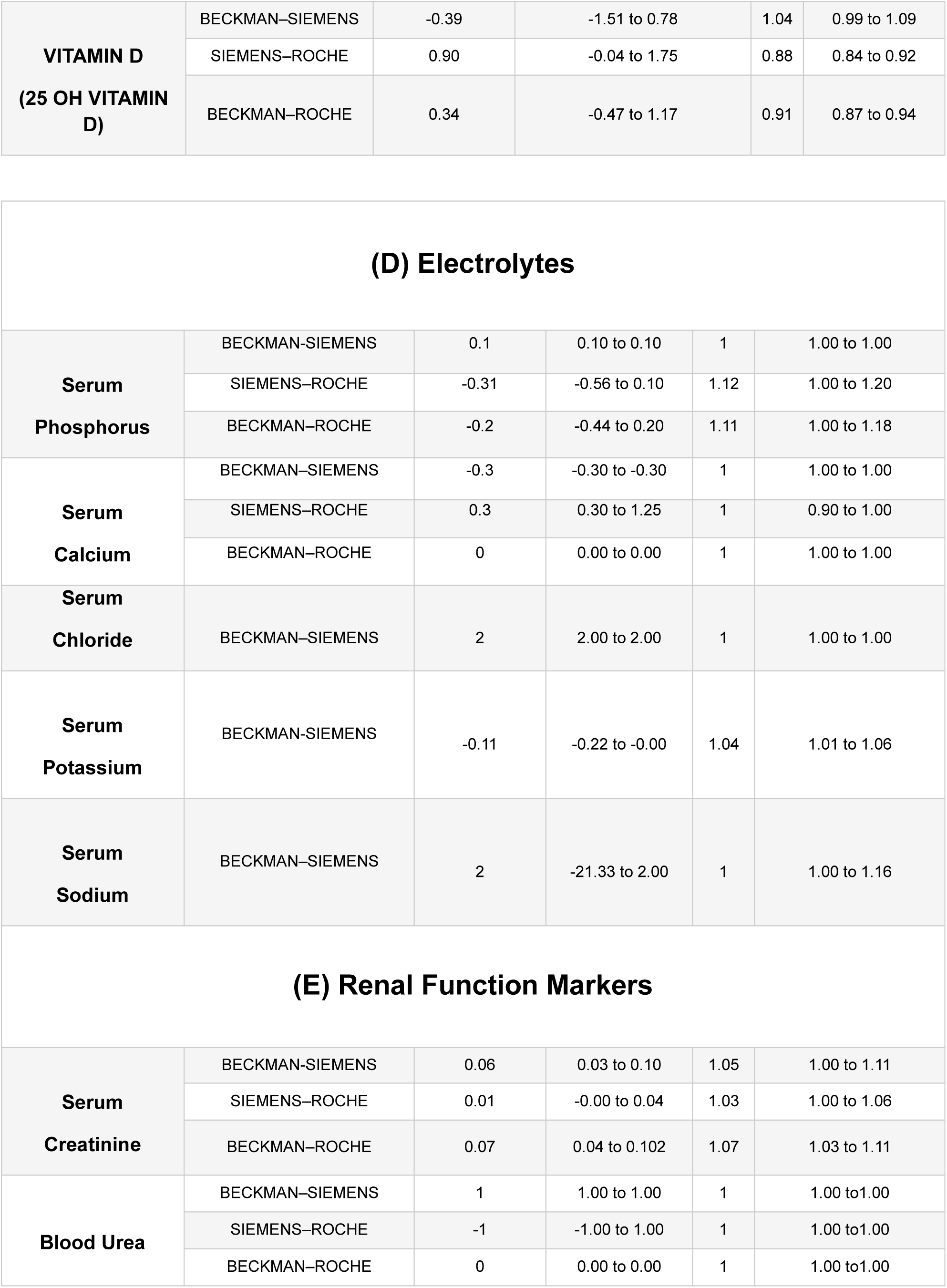
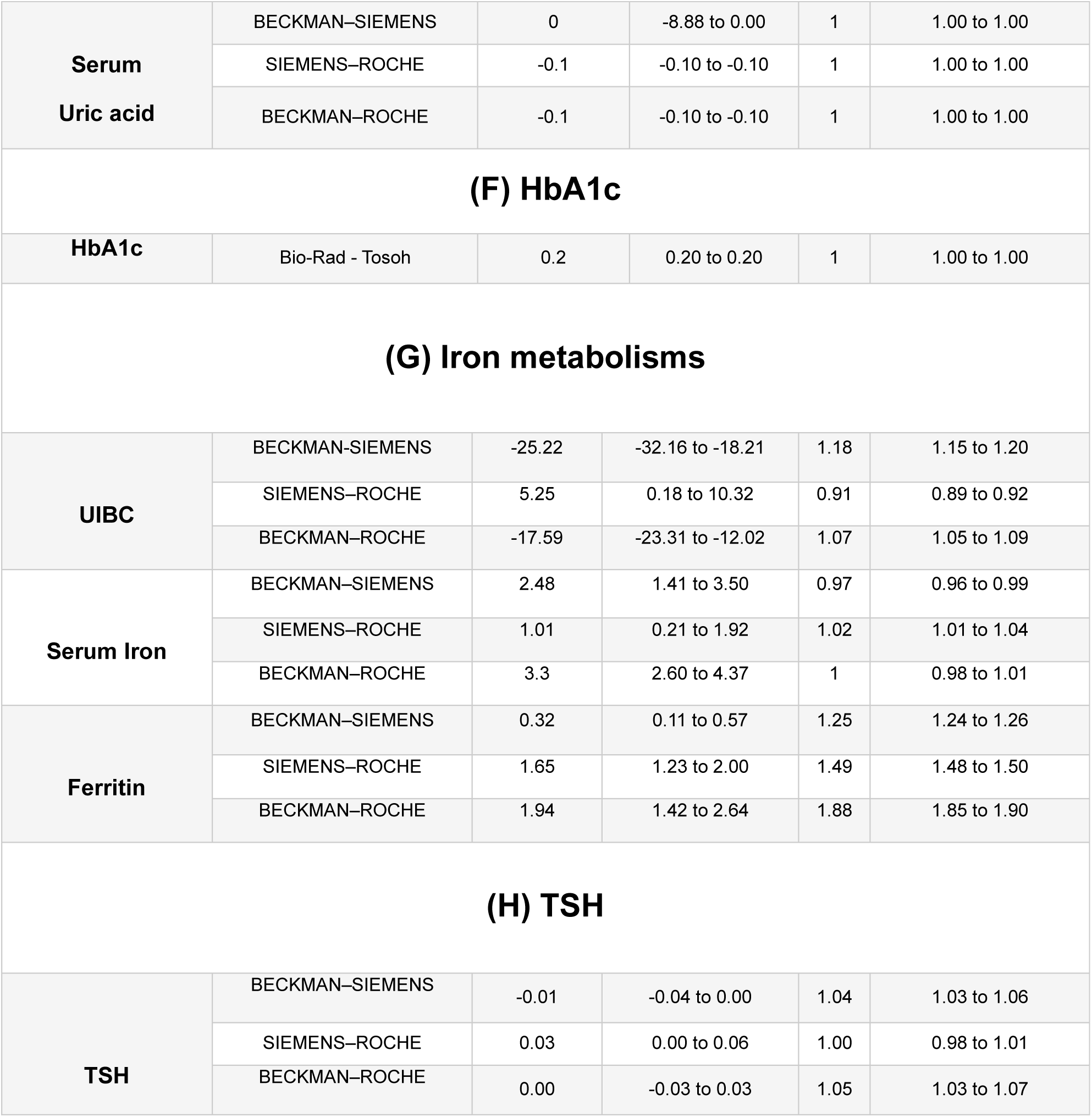
Passing Bablok Regression results.

**Table S2:**
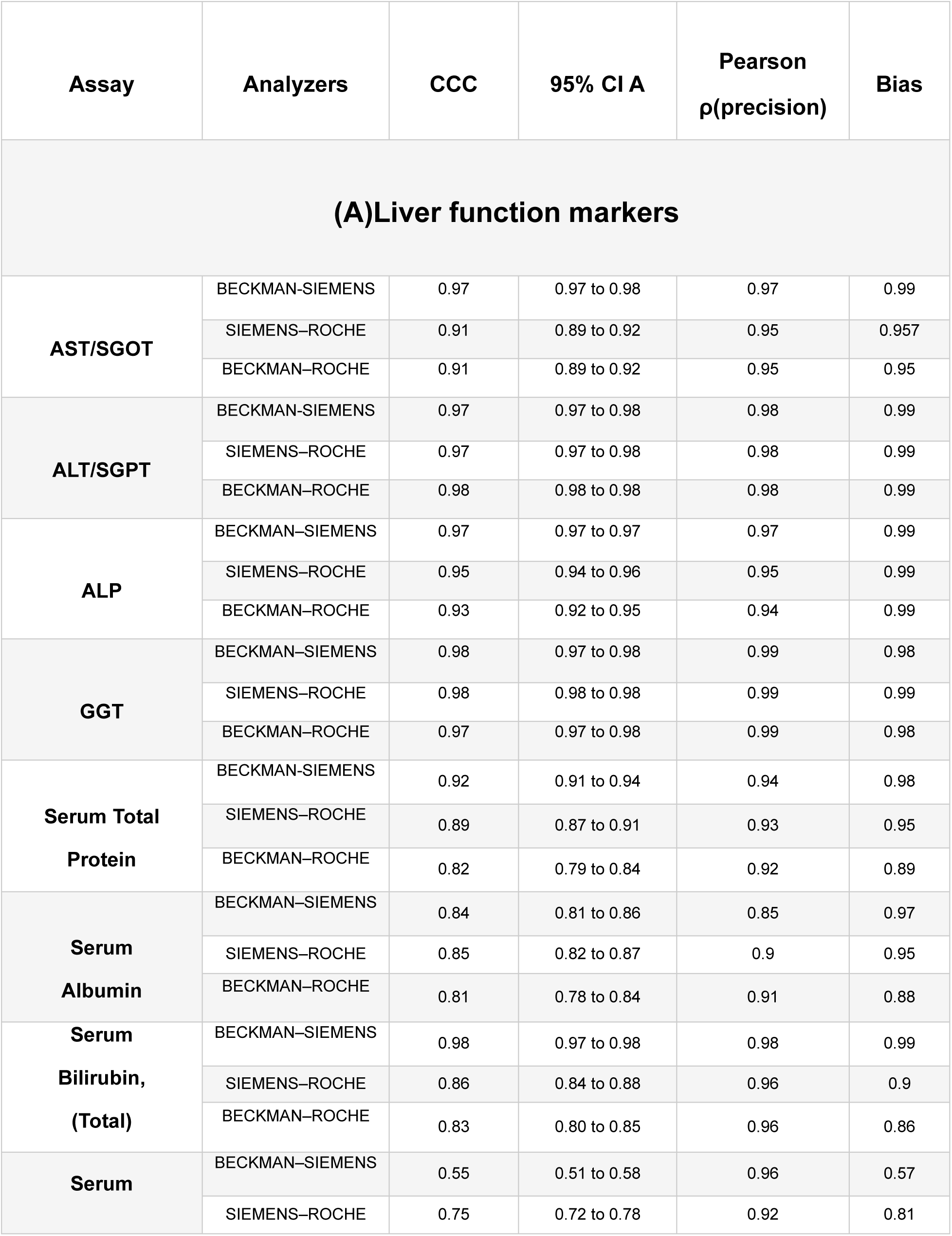
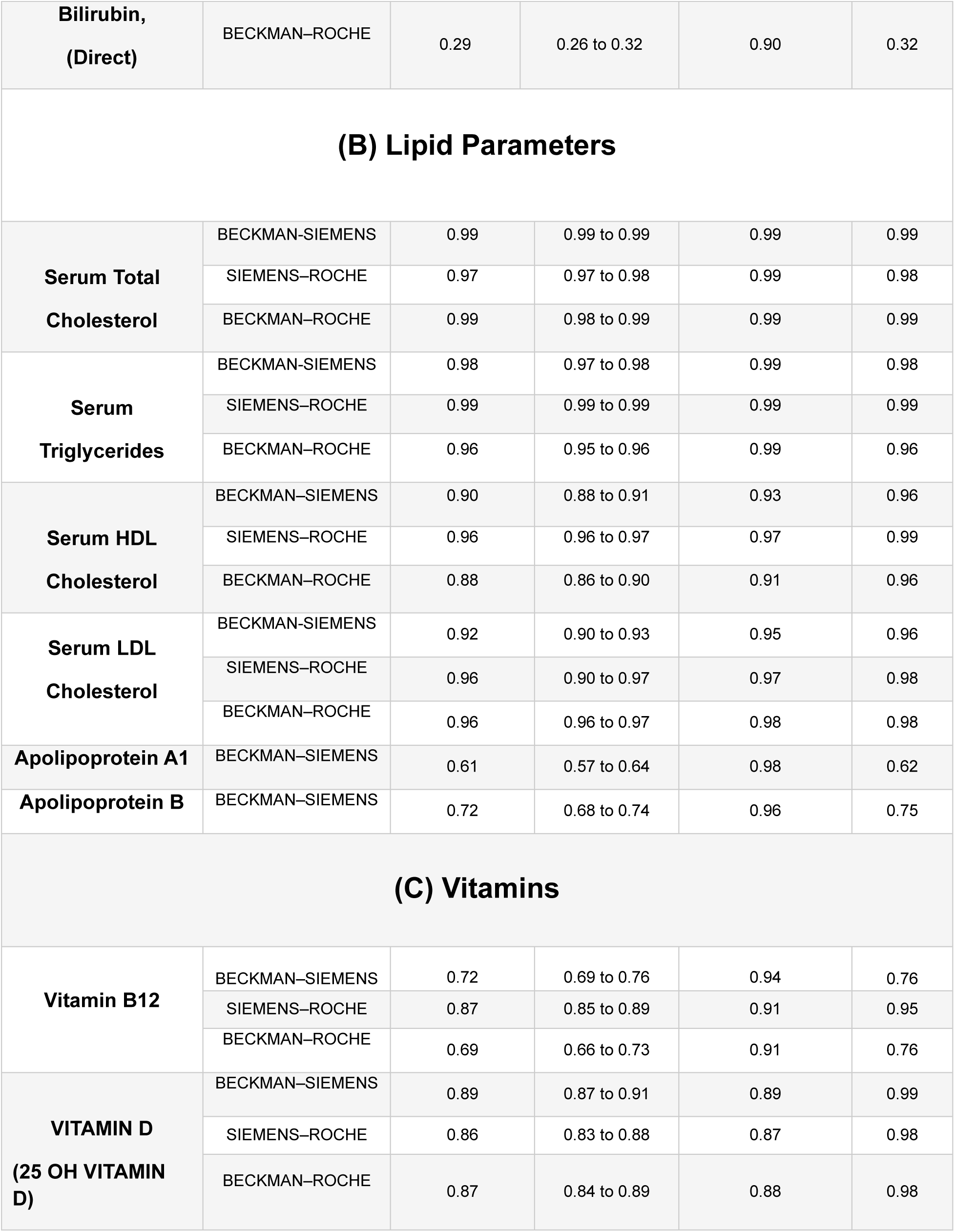
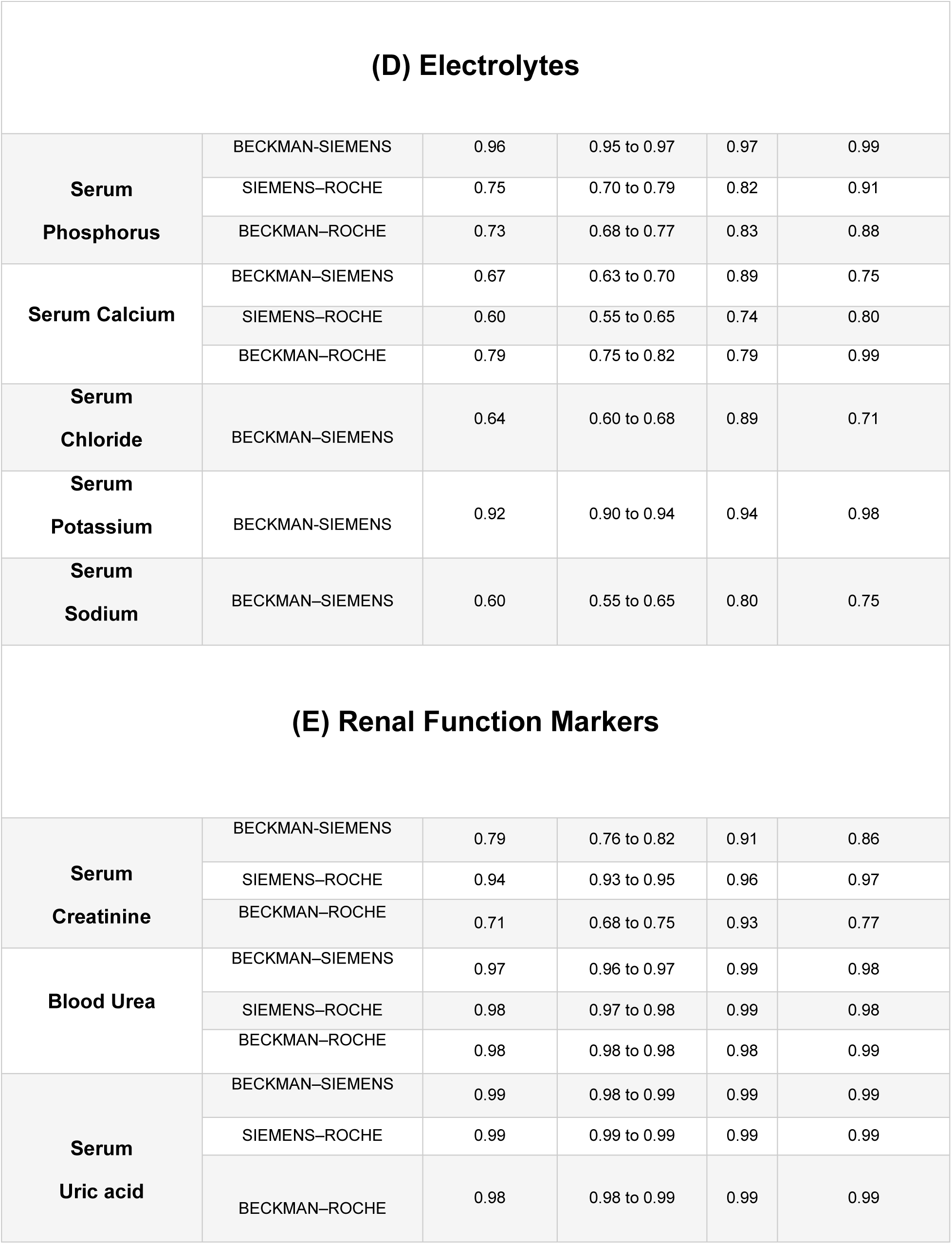
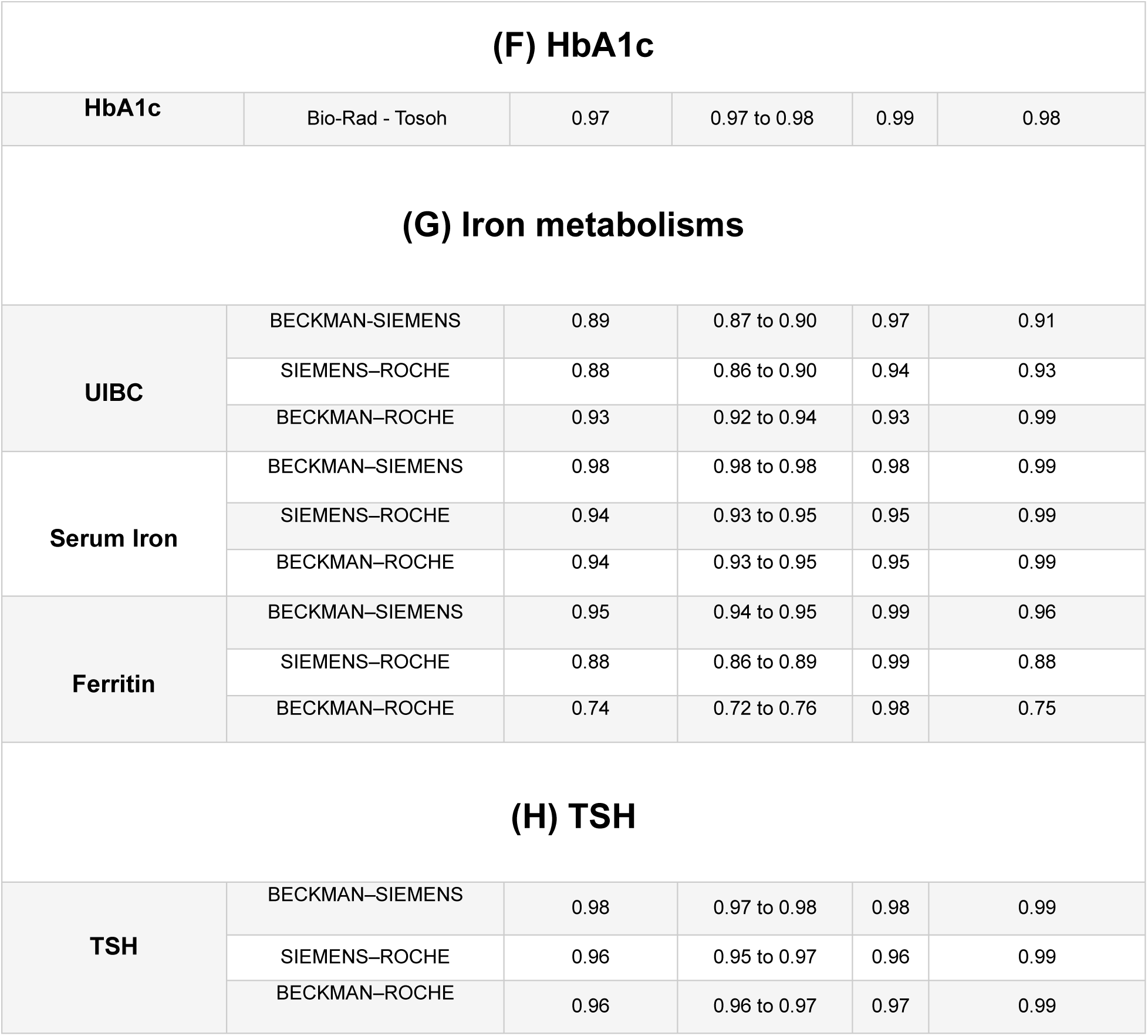
Concordance Correlation Coefficient results.

**Table S3:**
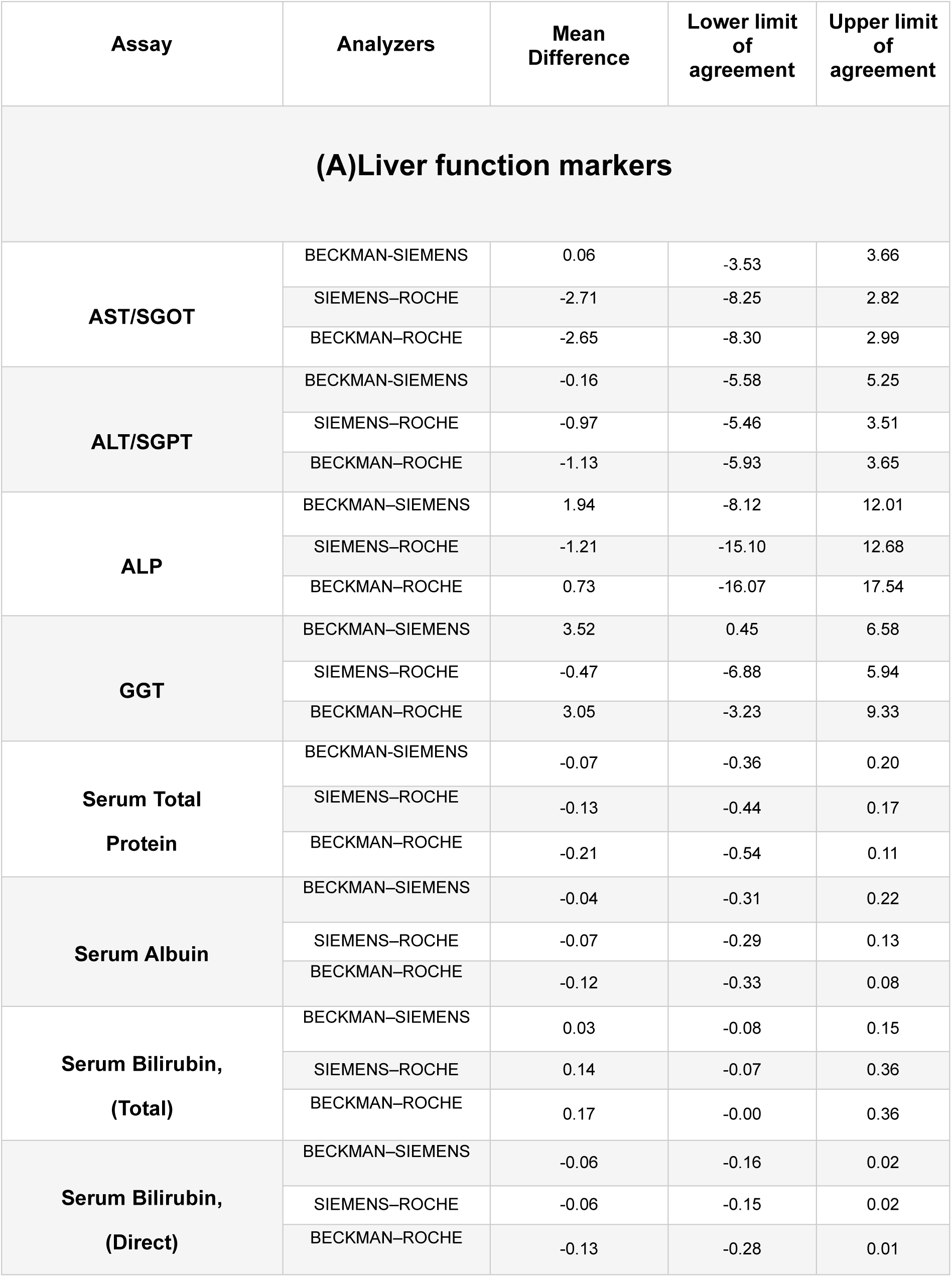
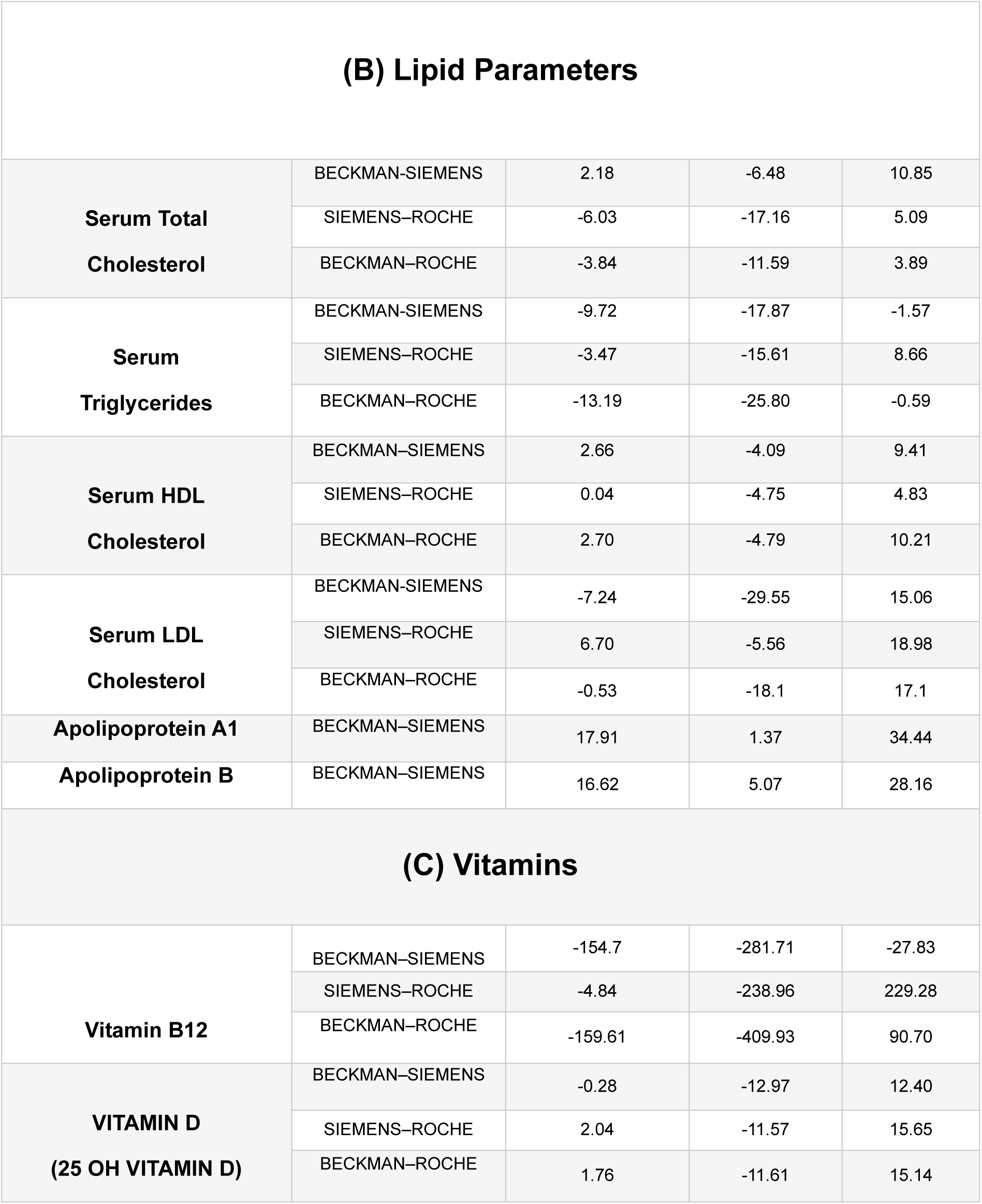
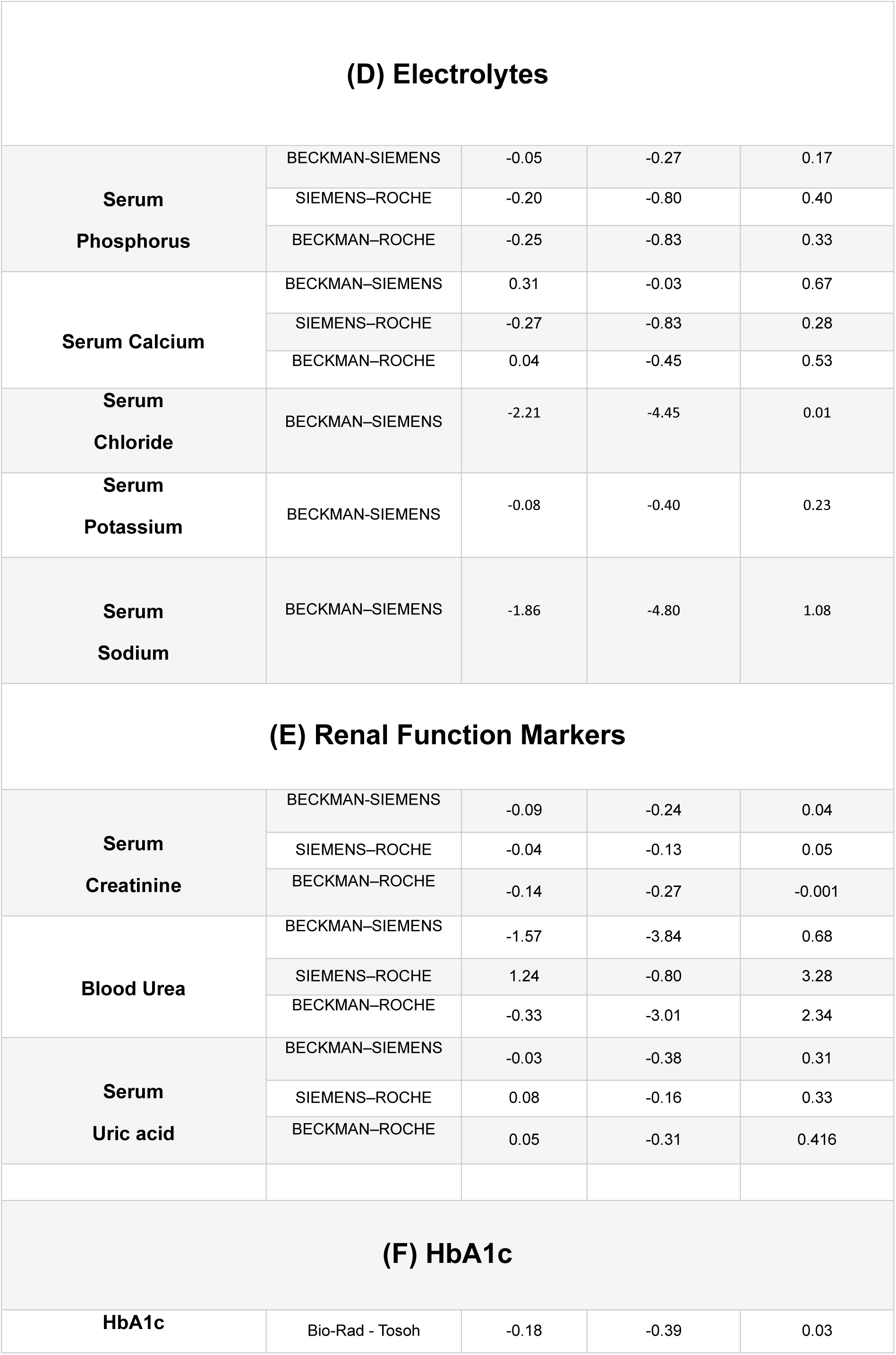
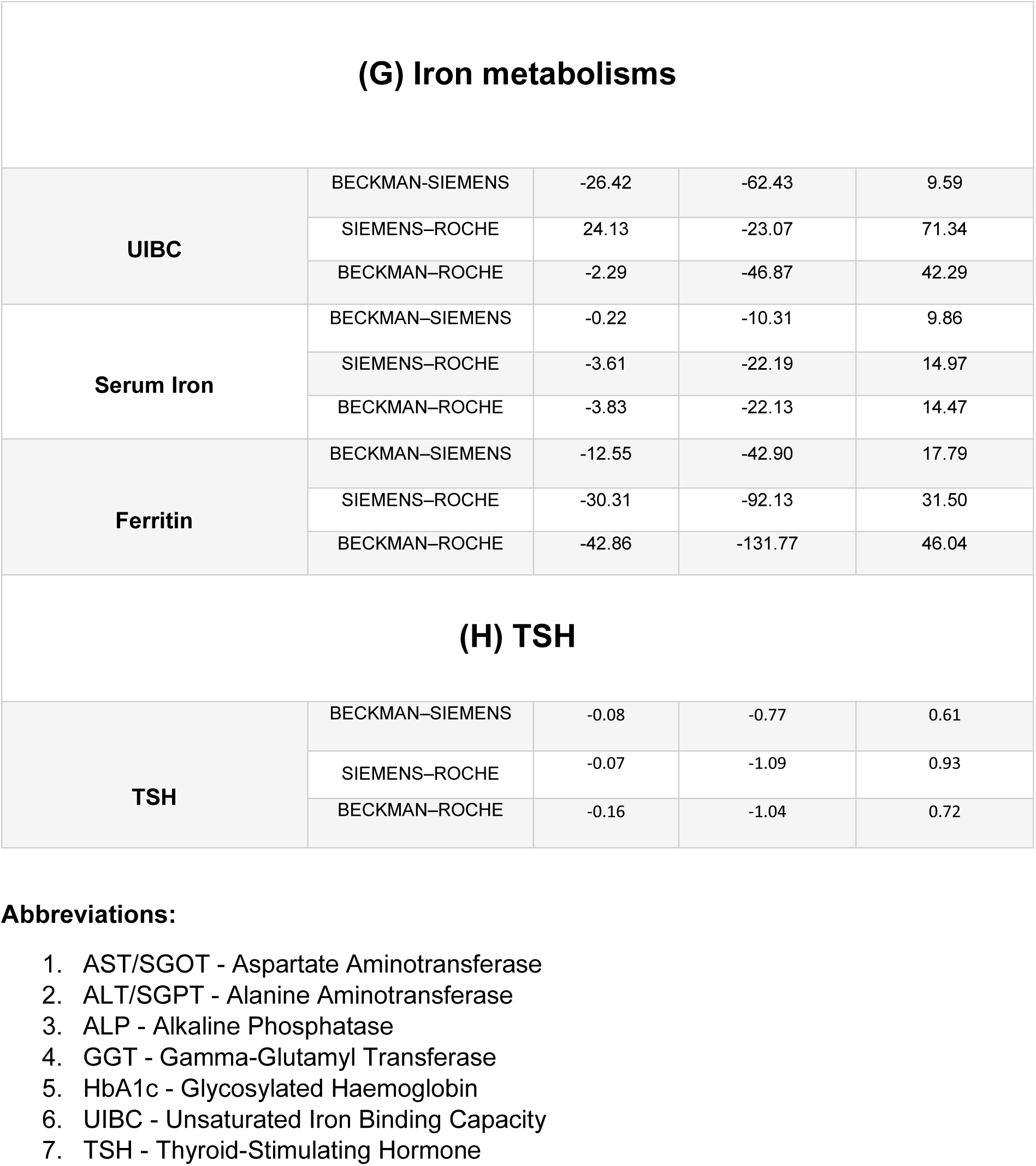
Bland Altman Analysis results.

### Supplementary Figures

**Figure S1:**
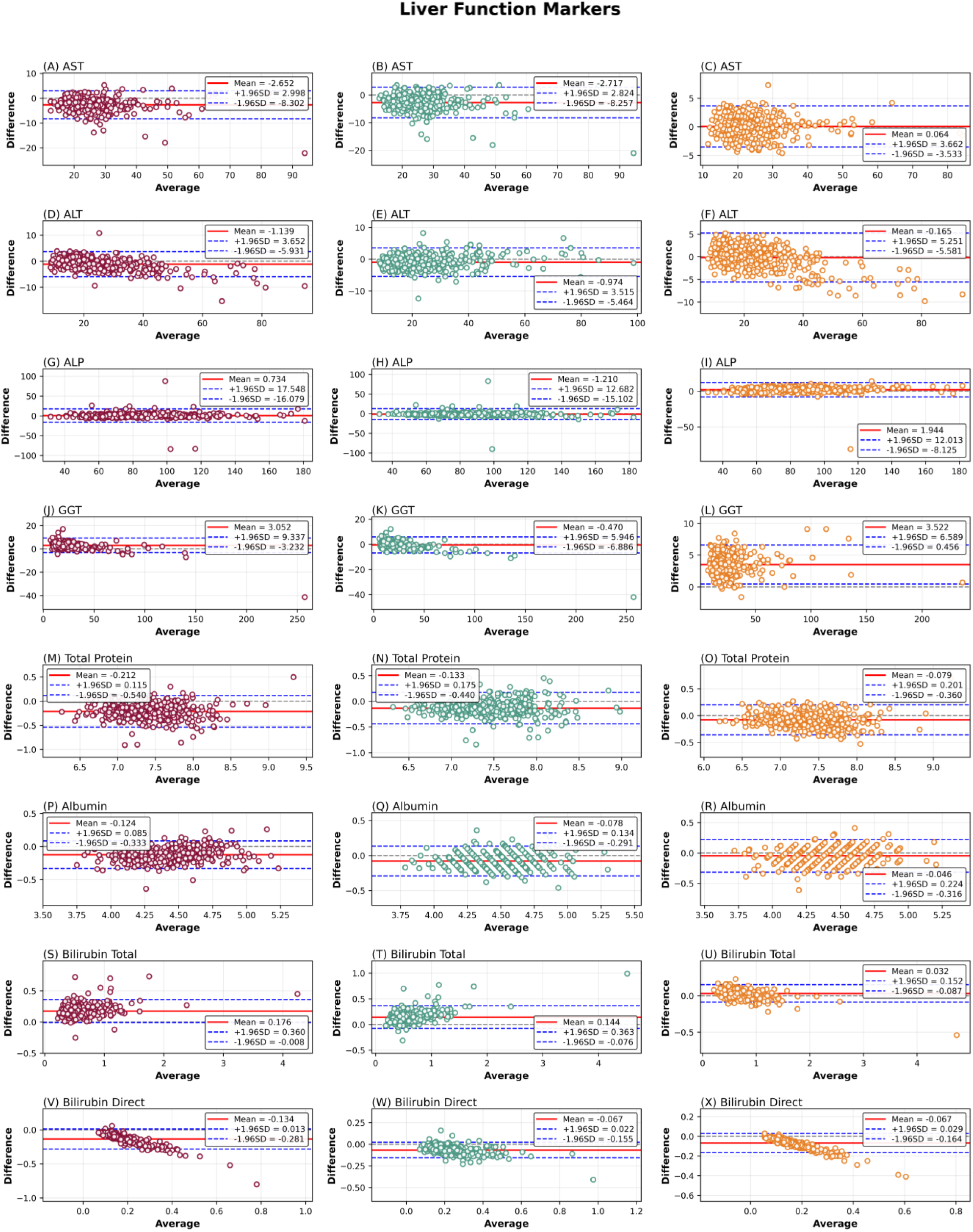
Bland-Altman analysis of agreement between clinical chemistry analyzers for liver function markers. Bland-Altman plots display the mean bias and 95% limits of agreement for AST(A-C), ALT (D-F), ALP(G-I), GGT(J-L), Total protein (M-O), Albumin(P-R), Bilirubin total (S-U), and Bilirubin direct (V-X) measured across different analyzers. The solid horizontal line represents the mean difference (bias), while the dashed lines indicate the upper and lower limits of agreement (mean ± 1.96 SD). Comparisons are shown for three analyzer pairings: Beckman-Roche (maroon), Siemens-Roche (green) and Beckman-Siemens (orange).

**Figure S2:**
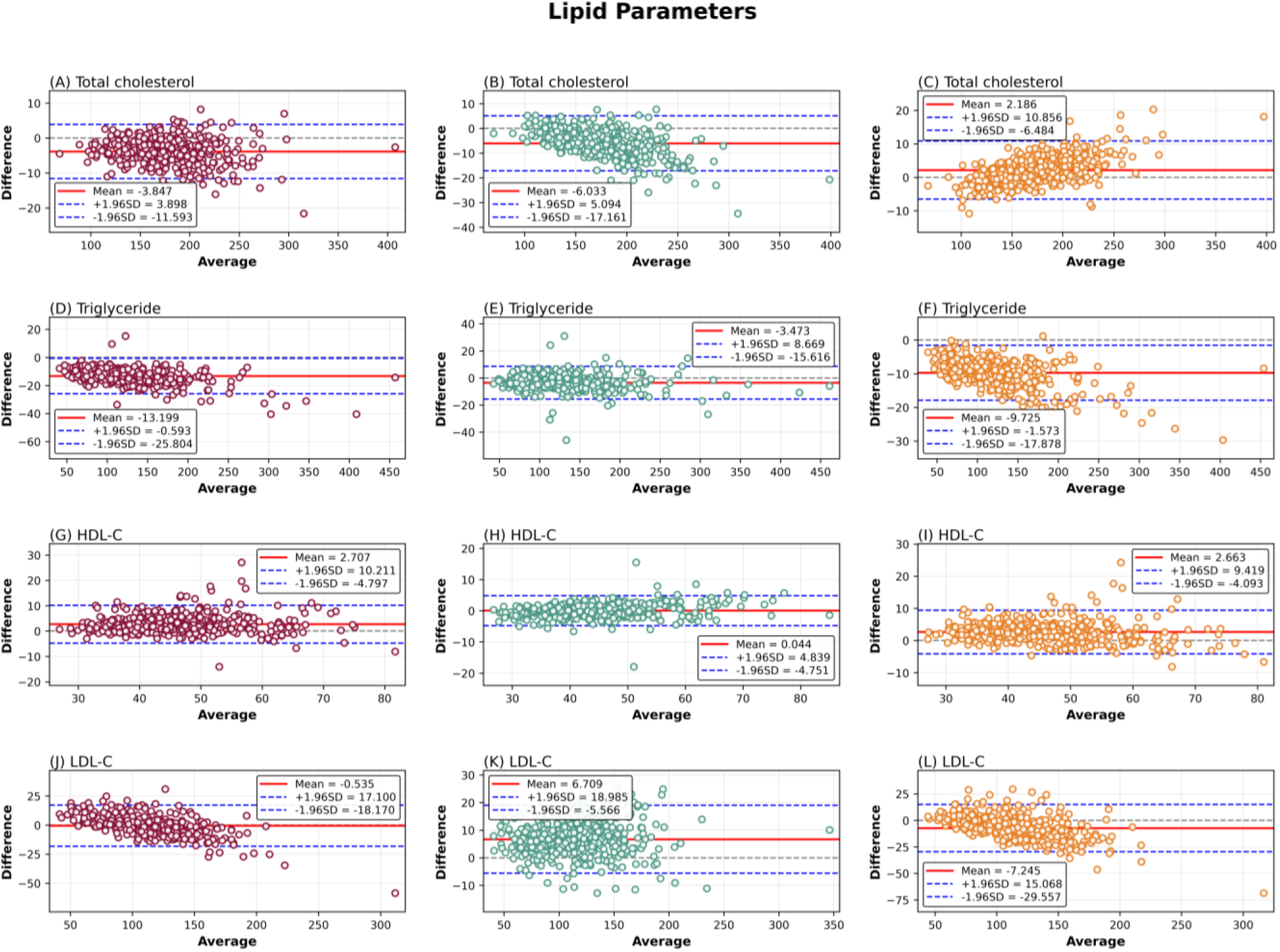
Bland-Altman analysis of agreement between clinical chemistry analyzers for lipid parameters. Bland-Altman plots illustrate the mean bias and 95% limits of agreement for Total cholesterol (A-C), Triglycerides (D-F), HDL-C (G-I) and LDL-C (J-L). The central horizontal line denotes the mean difference, and dashed lines represent the 95% limits of agreement (mean ± 1.96 SD). Three analyzer pairings are shown: Beckman-Roche (maroon), Siemens-Roche (green) and Beckman-Siemens (orange).

**Figure S3:**
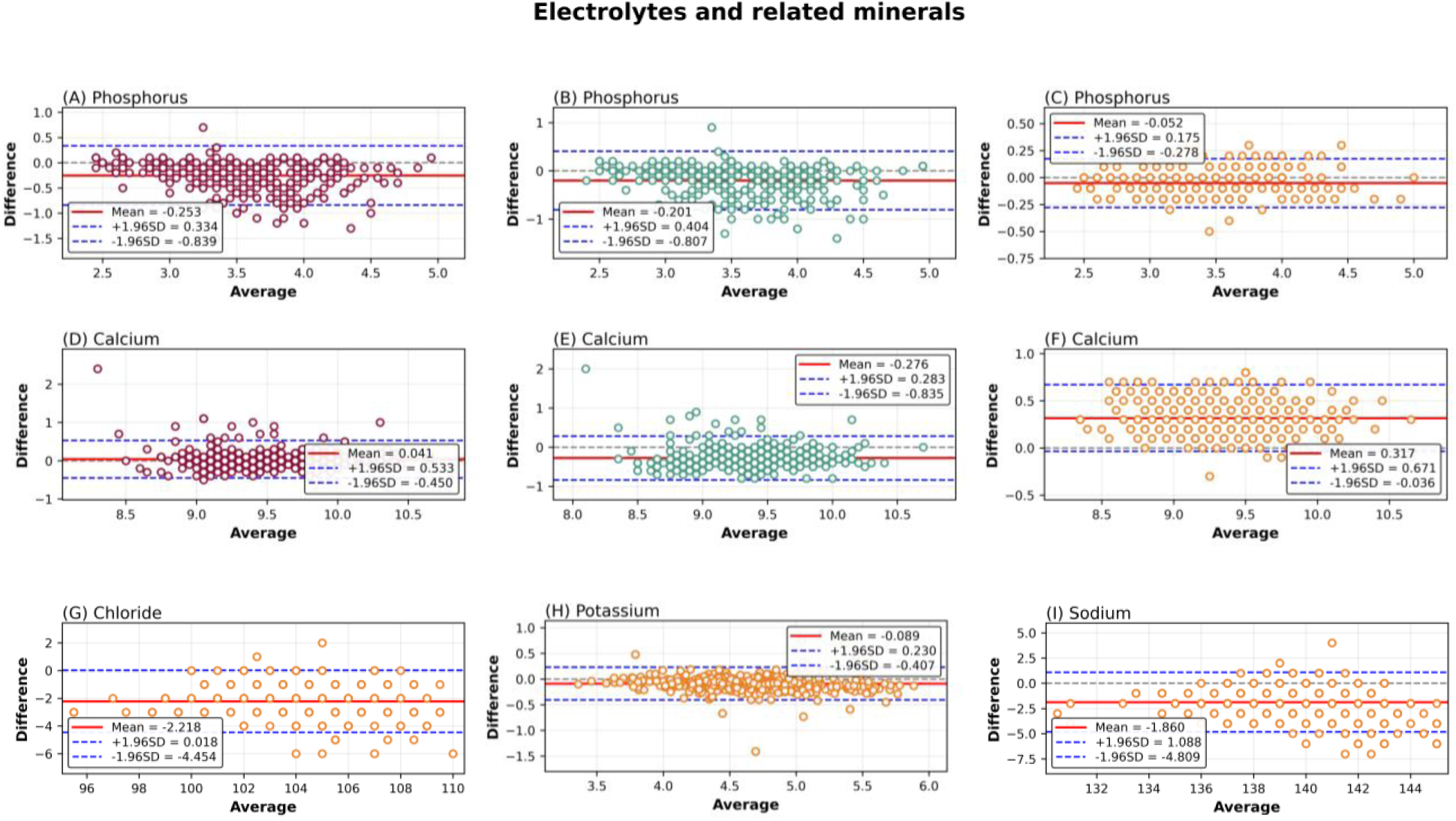
Bland-Altman analysis of agreement between clinical chemistry analyzers for electrolytes and related minerals. Bland-Altman plots depict the mean bias and 95% limits of agreement for Phosphorus (A-C), Calcium (D-F), Chloride (G), Potassium, (H) and Sodium (I) across different analyzers. The mean difference is indicated by the solid horizontal line, while dashed lines represent the upper and lower limits of agreement (mean ± 1.96 SD). Beckman-Roche (maroon), Siemens-Roche (green) and Beckman-Siemens (orange).

**Figure S4:**
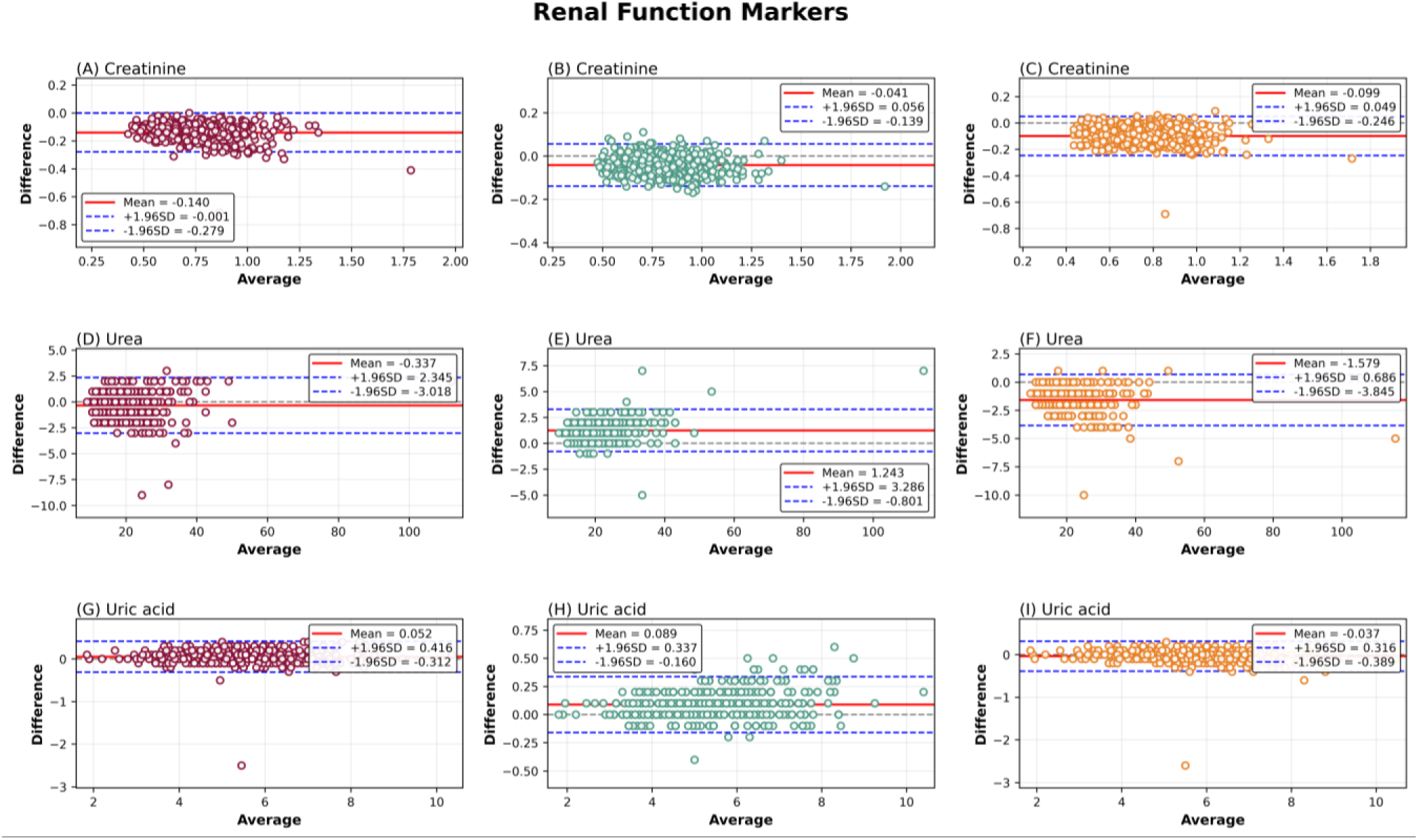
Bland-Altman analysis of agreement between clinical chemistry analyzers for renal function markers. Bland-Altman plots show the mean bias and 95% limits of agreement for Creatinine (A-C), Urea (D-F), and Uric acid (G-I) measured across analyzers. The solid horizontal line represents the mean difference, and the dashed lines correspond to the limits of agreement (mean ± 1.96 SD). Analyzer comparisons include Beckman-Roche (maroon), Siemens-Roche (green) and Beckman-Siemens (orange).

**Figure S5:**
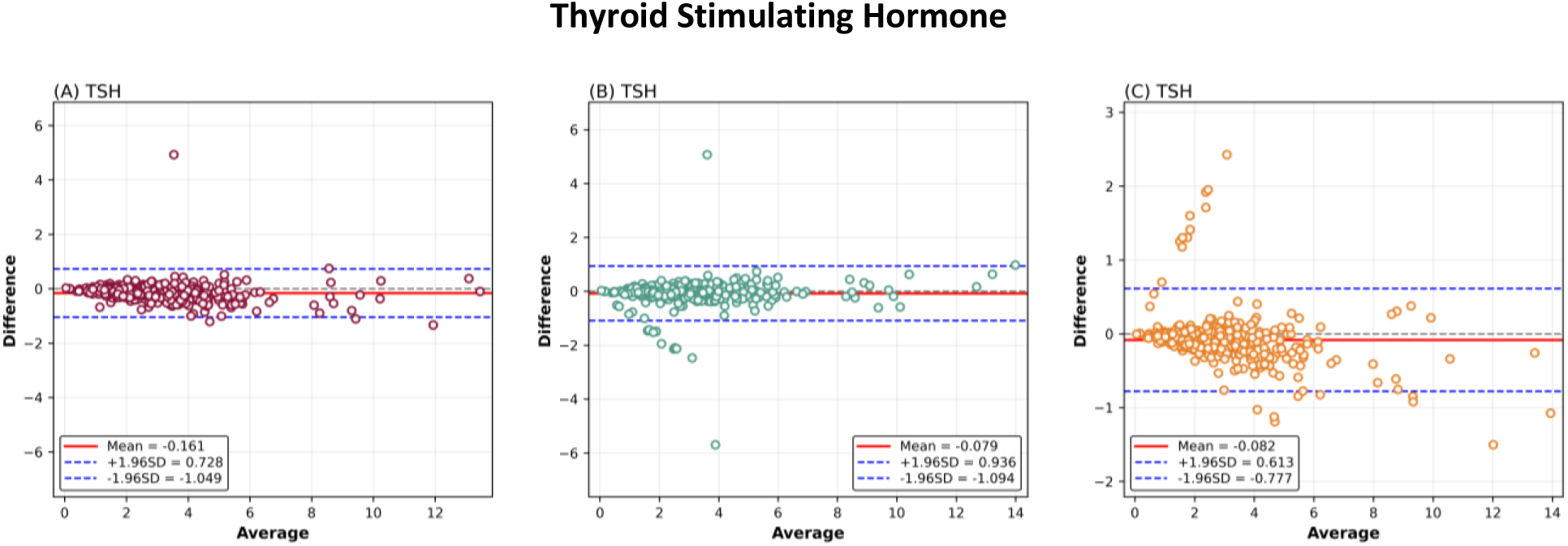
Bland-Altman analysis of agreement between immunoassay analyzers for TSH. Bland-Altman plots display the mean bias and 95% limits of agreement for Iron measurements across the compared analyzers. The solid horizontal line indicates the mean difference, while dashed lines denote the upper and lower limits of agreement (mean ± 1.96 SD). Analyzer comparisons include Beckman-Roche (maroon), Siemens-Roche (green) and Beckman-Siemens (orange).

**Figure S6:**
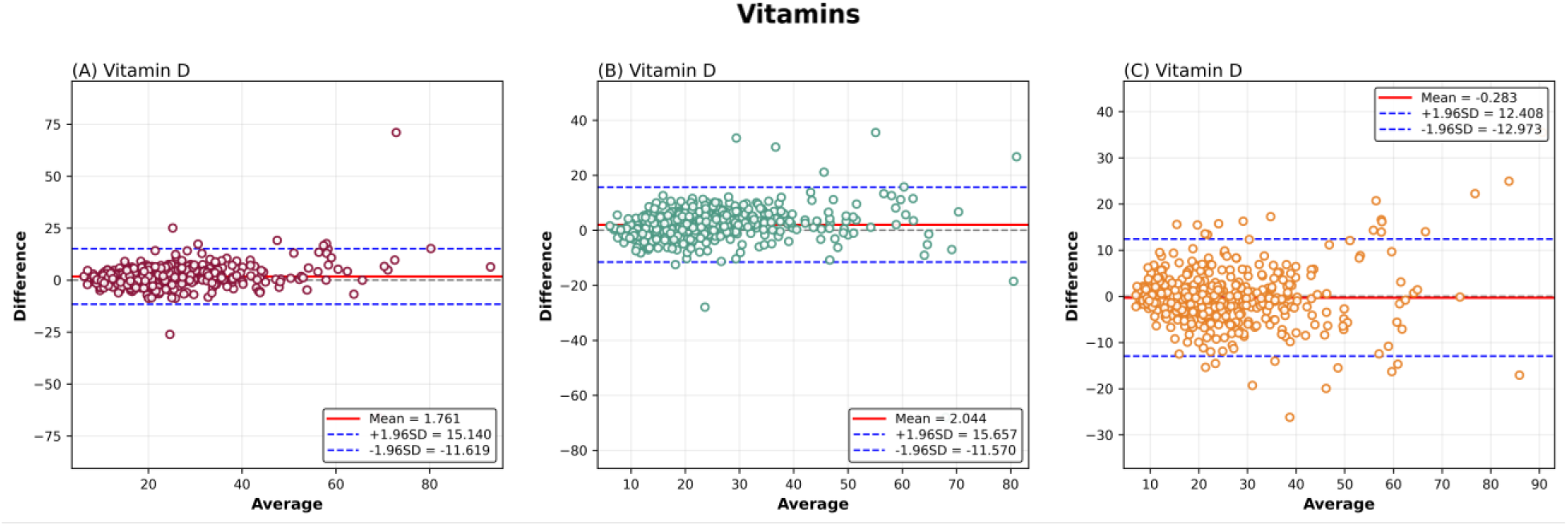
Bland-Altman analysis of agreement between immunoassay analyzers for vitamin measurements. Bland-Altman plots display the mean bias and 95% limits of agreement for Vitamin D concentrations across the compared analyzers. The solid horizontal line represents the mean difference, while dashed lines indicate the upper and lower limits of agreement (mean ± 1.96 SD). Analyzer comparisons include Beckman-Roche (maroon), Siemens-Roche (green) and Beckman-Siemens (orange).

**Figure S7:**
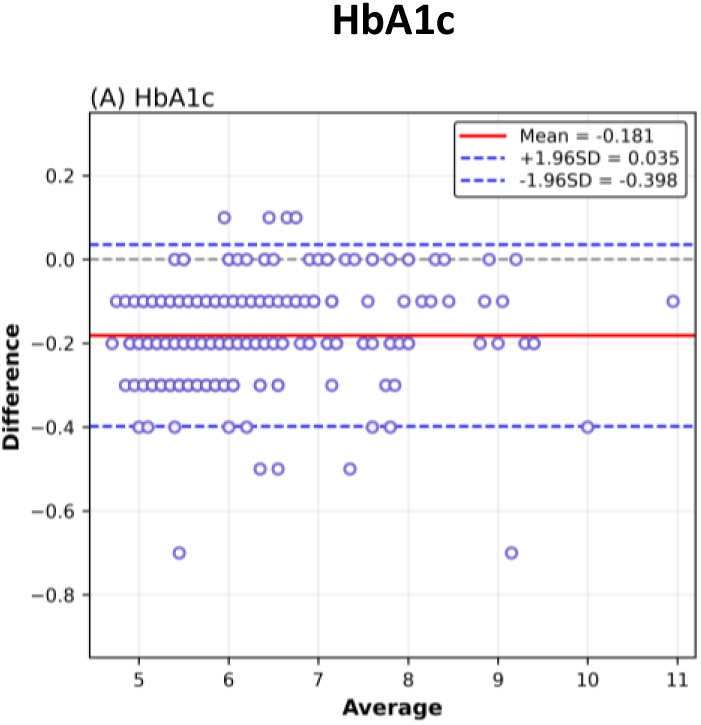
Bland-Altman analysis of agreement between analyzers for glycated haemoglobin (HbA1c). Bland-Altman plots present the mean bias and 95% limits of agreement for HbA1c values measured across different platforms. The solid line indicates the mean difference, and dashed lines represent the limits of agreement (mean ± 1.96 SD). Analyzer comparisons include Bio-rad - Tosoh.

