## Supplementary Material for "Assessing Inter-platform Variability of Blood Parameters across Three Automated Platforms: Report from Multi-centric Phenome India Study"

#### Supplementary Tables:

#### Table S1: Passing Bablok Regression results

| Assay | Analyzers | Intercept A | 95% CI A | Slope B | 95% CI B |
|---|---|---|---|---|---|
| **(A) Liver function markers** | | | | | |
| **AST/SGOT** | BECKMAN-SIEMENS | -0.21 | -0.87 to 0.36 | 1.00 | 0.98 to 1.02 |
| | SIEMENS–ROCHE | 2.19 | 1.43 to 2.96 | 1.02 | 0.98 to 1.05 |
| | BECKMAN–ROCHE | 1.87 | 1.03 to 2.73 | 1.02 | 0.99 to 1.06 |
| **ALT/SGPT** | BECKMAN-SIEMENS | -3.02 | -3.61 to -2.52 | 1.11 | 1.09 to 1.14 |
| | SIEMENS–ROCHE | 1.71 | 1.33 to 2.06 | 0.97 | 0.95 to 0.99 |
| | BECKMAN–ROCHE | -1.20 | -1.66 to -0.66 | 1.09 | 1.06 to 1.11 |
| **ALP** | BECKMAN-SIEMENS | 0.09 | -1.01 to 1.18 | 0.97 | 0.96 to 0.98 |
| | SIEMENS–ROCHE | 1.30 | 0.19 to 2.02 | 1.00 | 0.99 to 1.01 |
| | BECKMAN–ROCHE | 1.33 | -0.01 to 2.70 | 0.97 | 0.96 to 0.99 |
| **GGT** | BECKMAN-SIEMENS | -3.06 | -3.46 to -2.71 | 0.98 | 0.96 to 0.99 |
| | SIEMENS–ROCHE | -1.47 | -1.90 to -1.00 | 1.08 | 1.06 to 1.10 |
| | BECKMAN–ROCHE | -4.84 | -5.33 to -4.35 | 1.06 | 1.04 to 1.08 |
| **Serum Total Protein** | BECKMAN-SIEMENS | -0.28 | -0.55 to -0.02 | 1.05 | 1.01 to 1.08 |
| | SIEMENS–ROCHE | 0.31 | 0.14 to 0.54 | 0.97 | 0.94 to 1.00 |
| | BECKMAN–ROCHE | 0.05 | -0.24 to 0.31 | 1.02 | 0.98 to 1.06 |

| | | | | | |
|---|---|---|---|---|---|
| Serum Albumin | BECKMAN-SIEMENS | 0.52 | 0.31 to 0.76 | 0.88 | 0.83 to 0.93 |
| | SIEMENS–ROCHE | 0.08 | -0.09 to 0.30 | 1 | 0.95 to 1.04 |
| | BECKMAN–ROCHE | 0.60 | 0.43 to 0.76 | 0.88 | 0.85 to 0.92 |
| Serum Bilirubin, (Total) | BECKMAN-SIEMENS | -0.85 | -0.09 to -0.07 | 1.07 | 1.05 to 1.09 |
| | SIEMENS–ROCHE | -0.03 | -0.05 to -0.02 | 0.84 | 0.82 to 0.87 |
| | BECKMAN–ROCHE | -0.11 | -0.13 to -0.09 | 0.92 | 0.90 to 0.95 |
| Serum Bilirubin, (Direct) | BECKMAN-SIEMENS | -0.5 | -0.05 to -0.03 | 2 | 1.87 to 2.00 |
| | SIEMENS–ROCHE | 0.02 | 0.01 to 0.03 | 1.26 | 1.21 to 1.33 |
| | BECKMAN–ROCHE | -0.04 | -0.05 to -0.03 | 2.5 | 2.40 to 2.62 |

### (B) Lipid Parameters

| | | | | | |
|---|---|---|---|---|---|
| Serum Total Cholesterol | BECKMAN-SIEMENS | 8.60 | 7.12 to 10.09 | 0.93 | 0.93 to 0.94 |
| | SIEMENS–ROCHE | -7.91 | -10.27 to -5.75 | 1.08 | 1.06 to 1.09 |
| | BECKMAN–ROCHE | 1.43 | -0.16 to 2.93 | 1.01 | 1.00 to 1.02 |
| Serum Triglycerides | BECKMAN-SIEMENS | 4.53 | 3.64 to 5.42 | 1.04 | 1.03 to 1.05 |
| | SIEMENS–ROCHE | 2.22 | 1.02 to 3.50 | 1.01 | 1.00 to 1.02 |
| | BECKMAN–ROCHE | 6.44 | 5.18 to 7.58 | 1.05 | 1.04 to 1.06 |
| Serum HDL Cholesterol | BECKMAN–SIEMENS | -5.86 | -7.44 to -4.23 | 1.07 | 1.03 to 1.10 |
| | SIEMENS–ROCHE | 2.60 | 1.66 to 3.58 | 0.93 | 0.91 to 0.95 |
| | BECKMAN–ROCHE | -2.1 | -3.86 to -0.57 | 1 | 0.96 to 1.03 |
| Serum LDL Cholesterol | BECKMAN-SIEMENS | -15.59 | -18.96 to -12.27 | 1.21 | 1.18 to1.24 |
| | SIEMENS–ROCHE | -4.12 | -6.06 to -2.142 | 0.97 | 0.96to0.99 |
| | BECKMAN–ROCHE | -19.21 | -21.93 to -16.65 | 1.17 | 1.15to1.20 |
| Apolipoprotein A1 | BECKMAN–SIEMENS | 25.81 | 24.15 to 27.48 | 0.65 | 0.64 to 0.67 |
| Apolipoprotein B | BECKMAN–SIEMENS | -12.43 | -14.86 to -9.79 | 0.96 | 0.93 to 0.99 |

| (C) Vitamins | | | | | |
|---|---|---|---|---|---|
| Vitamin B12 | BECKMAN–SIEMENS | 133.63 | 126.52 to 139.92 | 1.11 | 1.07 to 1.15 |
| | SIEMENS–ROCHE | -142.99 | -160.07 to -127.97 | 1.36 | 1.31 to 1.41 |
| | BECKMAN–ROCHE | 34.40 | 27.03 to 41.10 | 1.57 | 1.54 to 1.61 |
| VITAMIN D (25 OH VITAMIN D) | BECKMAN–SIEMENS | -0.39 | -1.51 to 0.78 | 1.04 | 0.99 to 1.09 |
| | SIEMENS–ROCHE | 0.90 | -0.04 to 1.75 | 0.88 | 0.84 to 0.92 |
| | BECKMAN–ROCHE | 0.34 | -0.47 to 1.17 | 0.91 | 0.87 to 0.94 |

| (D) Electrolytes | | | | | |
|---|---|---|---|---|---|
| Serum Phosphorus | BECKMAN-SIEMENS | 0.1 | 0.10 to 0.10 | 1 | 1.00 to 1.00 |
| | SIEMENS–ROCHE | -0.31 | -0.56 to 0.10 | 1.12 | 1.00 to 1.20 |
| | BECKMAN–ROCHE | -0.2 | -0.44 to 0.20 | 1.11 | 1.00 to 1.18 |
| Serum Calcium | BECKMAN–SIEMENS | -0.3 | -0.30 to -0.30 | 1 | 1.00 to 1.00 |
| | SIEMENS–ROCHE | 0.3 | 0.30 to 1.25 | 1 | 0.90 to 1.00 |
| | BECKMAN–ROCHE | 0 | 0.00 to 0.00 | 1 | 1.00 to 1.00 |
| Serum Chloride | BECKMAN–SIEMENS | 2 | 2.00 to 2.00 | 1 | 1.00 to 1.00 |
| Serum Potassium | BECKMAN-SIEMENS | -0.11 | -0.22 to -0.00 | 1.04 | 1.01 to 1.06 |
| Serum Sodium | BECKMAN–SIEMENS | 2 | -21.33 to 2.00 | 1 | 1.00 to 1.16 |
| (E) Renal Function Markers | | | | | |

| | | | | | |
|---|---|---|---|---|---|
| Serum Creatinine | BECKMAN-SIEMENS | 0.06 | 0.03 to 0.10 | 1.05 | 1.00 to 1.11 |
| | SIEMENS–ROCHE | 0.01 | -0.00 to 0.04 | 1.03 | 1.00 to 1.06 |
| | BECKMAN–ROCHE | 0.07 | 0.04 to 0.102 | 1.07 | 1.03 to 1.11 |
| Blood Urea | BECKMAN–SIEMENS | 1 | 1.00 to 1.00 | 1 | 1.00 to1.00 |
| | SIEMENS–ROCHE | -1 | -1.00 to 1.00 | 1 | 1.00 to1.00 |
| | BECKMAN–ROCHE | 0 | 0.00 to 0.00 | 1 | 1.00 to1.00 |
| Serum Uric acid | BECKMAN–SIEMENS | 0 | -8.88 to 0.00 | 1 | 1.00 to 1.00 |
| | SIEMENS–ROCHE | -0.1 | -0.10 to -0.10 | 1 | 1.00 to 1.00 |
| | BECKMAN–ROCHE | -0.1 | -0.10 to -0.10 | 1 | 1.00 to 1.00 |
| **(F) HbA1c** | | | | | |
| HbA1c | Bio-Rad - Tosoh | 0.2 | 0.20 to 0.20 | 1 | 1.00 to 1.00 |
| **(G) Iron metabolisms** | | | | | |
| UIBC | BECKMAN-SIEMENS | -25.22 | -32.16 to -18.21 | 1.18 | 1.15 to 1.20 |
| | SIEMENS–ROCHE | 5.25 | 0.18 to 10.32 | 0.91 | 0.89 to 0.92 |
| | BECKMAN–ROCHE | -17.59 | -23.31 to -12.02 | 1.07 | 1.05 to 1.09 |
| Serum Iron | BECKMAN–SIEMENS | 2.48 | 1.41 to 3.50 | 0.97 | 0.96 to 0.99 |
| | SIEMENS–ROCHE | 1.01 | 0.21 to 1.92 | 1.02 | 1.01 to 1.04 |
| | BECKMAN–ROCHE | 3.3 | 2.60 to 4.37 | 1 | 0.98 to 1.01 |
| Ferritin | BECKMAN–SIEMENS | 0.32 | 0.11 to 0.57 | 1.25 | 1.24 to 1.26 |
| | SIEMENS–ROCHE | 1.65 | 1.23 to 2.00 | 1.49 | 1.48 to 1.50 |
| | BECKMAN–ROCHE | 1.94 | 1.42 to 2.64 | 1.88 | 1.85 to 1.90 |
| **(H) TSH** | | | | | |
| TSH | BECKMAN–SIEMENS | -0.01 | -0.04 to 0.00 | 1.04 | 1.03 to 1.06 |
| | SIEMENS–ROCHE | 0.03 | 0.00 to 0.06 | 1.00 | 0.98 to 1.01 |
| | BECKMAN–ROCHE | 0.00 | -0.03 to 0.03 | 1.05 | 1.03 to 1.07 |

**Table S2: Concordance Correlation Coefficient results**

| Assay | Analyzers | CCC | 95% CI A | Pearson ρ(precision) | Bias |
|---|---|---|---|---|---|
| **(A)Liver function markers** | | | | | |
| **AST/SGOT** | BECKMAN-SIEMENS | 0.97 | 0.97 to 0.98 | 0.97 | 0.99 |
| | SIEMENS–ROCHE | 0.91 | 0.89 to 0.92 | 0.95 | 0.957 |
| | BECKMAN–ROCHE | 0.91 | 0.89 to 0.92 | 0.95 | 0.95 |
| **ALT/SGPT** | BECKMAN-SIEMENS | 0.97 | 0.97 to 0.98 | 0.98 | 0.99 |
| | SIEMENS–ROCHE | 0.97 | 0.97 to 0.98 | 0.98 | 0.99 |
| | BECKMAN–ROCHE | 0.98 | 0.98 to 0.98 | 0.98 | 0.99 |
| **ALP** | BECKMAN–SIEMENS | 0.97 | 0.97 to 0.97 | 0.97 | 0.99 |
| | SIEMENS–ROCHE | 0.95 | 0.94 to 0.96 | 0.95 | 0.99 |
| | BECKMAN–ROCHE | 0.93 | 0.92 to 0.95 | 0.94 | 0.99 |
| **GGT** | BECKMAN–SIEMENS | 0.98 | 0.97 to 0.98 | 0.99 | 0.98 |
| | SIEMENS–ROCHE | 0.98 | 0.98 to 0.98 | 0.99 | 0.99 |
| | BECKMAN–ROCHE | 0.97 | 0.97 to 0.98 | 0.99 | 0.98 |
| **Serum Total Protein** | BECKMAN-SIEMENS | 0.92 | 0.91 to 0.94 | 0.94 | 0.98 |
| | SIEMENS–ROCHE | 0.89 | 0.87 to 0.91 | 0.93 | 0.95 |
| | BECKMAN–ROCHE | 0.82 | 0.79 to 0.84 | 0.92 | 0.89 |
| **Serum Albumin** | BECKMAN–SIEMENS | 0.84 | 0.81 to 0.86 | 0.85 | 0.97 |
| | SIEMENS–ROCHE | 0.85 | 0.82 to 0.87 | 0.9 | 0.95 |
| | BECKMAN–ROCHE | 0.81 | 0.78 to 0.84 | 0.91 | 0.88 |
| **Serum Bilirubin, (Total)** | BECKMAN–SIEMENS | 0.98 | 0.97 to 0.98 | 0.98 | 0.99 |
| | SIEMENS–ROCHE | 0.86 | 0.84 to 0.88 | 0.96 | 0.9 |
| | BECKMAN–ROCHE | 0.83 | 0.80 to 0.85 | 0.96 | 0.86 |
| **Serum** | BECKMAN–SIEMENS | 0.55 | 0.51 to 0.58 | 0.96 | 0.57 |
| | SIEMENS–ROCHE | 0.75 | 0.72 to 0.78 | 0.92 | 0.81 |

| | | | | | |
|---|---|---|---|---|---|
| **Bilirubin, (Direct)** | BECKMAN–ROCHE | 0.29 | 0.26 to 0.32 | 0.90 | 0.32 |
| **(B) Lipid Parameters** | | | | | |
| **Serum Total Cholesterol** | BECKMAN-SIEMENS | 0.99 | 0.99 to 0.99 | 0.99 | 0.99 |
| | SIEMENS–ROCHE | 0.97 | 0.97 to 0.98 | 0.99 | 0.98 |
| | BECKMAN–ROCHE | 0.99 | 0.98 to 0.99 | 0.99 | 0.99 |
| **Serum Triglycerides** | BECKMAN-SIEMENS | 0.98 | 0.97 to 0.98 | 0.99 | 0.98 |
| | SIEMENS–ROCHE | 0.99 | 0.99 to 0.99 | 0.99 | 0.99 |
| | BECKMAN–ROCHE | 0.96 | 0.95 to 0.96 | 0.99 | 0.96 |
| **Serum HDL Cholesterol** | BECKMAN–SIEMENS | 0.90 | 0.88 to 0.91 | 0.93 | 0.96 |
| | SIEMENS–ROCHE | 0.96 | 0.96 to 0.97 | 0.97 | 0.99 |
| | BECKMAN–ROCHE | 0.88 | 0.86 to 0.90 | 0.91 | 0.96 |
| **Serum LDL Cholesterol** | BECKMAN-SIEMENS | 0.92 | 0.90 to 0.93 | 0.95 | 0.96 |
| | SIEMENS–ROCHE | 0.96 | 0.90 to 0.97 | 0.97 | 0.98 |
| | BECKMAN–ROCHE | 0.96 | 0.96 to 0.97 | 0.98 | 0.98 |
| **Apolipoprotein A1** | BECKMAN–SIEMENS | 0.61 | 0.57 to 0.64 | 0.98 | 0.62 |
| **Apolipoprotein B** | BECKMAN–SIEMENS | 0.72 | 0.68 to 0.74 | 0.96 | 0.75 |
| **(C) Vitamins** | | | | | |
| **Vitamin B12** | BECKMAN–SIEMENS | 0.72 | 0.69 to 0.76 | 0.94 | 0.76 |
| | SIEMENS–ROCHE | 0.87 | 0.85 to 0.89 | 0.91 | 0.95 |
| | BECKMAN–ROCHE | 0.69 | 0.66 to 0.73 | 0.91 | 0.76 |
| **VITAMIN D (25 OH VITAMIN D)** | BECKMAN–SIEMENS | 0.89 | 0.87 to 0.91 | 0.89 | 0.99 |
| | SIEMENS–ROCHE | 0.86 | 0.83 to 0.88 | 0.87 | 0.98 |
| | BECKMAN–ROCHE | 0.87 | 0.84 to 0.89 | 0.88 | 0.98 |

| | | | | | |
|---|---|---|---|---|---|
| **(D) Electrolytes** | | | | | |
| **Serum Phosphorus** | BECKMAN-SIEMENS | 0.96 | 0.95 to 0.97 | 0.97 | 0.99 |
| | SIEMENS–ROCHE | 0.75 | 0.70 to 0.79 | 0.82 | 0.91 |
| | BECKMAN–ROCHE | 0.73 | 0.68 to 0.77 | 0.83 | 0.88 |
| **Serum Calcium** | BECKMAN–SIEMENS | 0.67 | 0.63 to 0.70 | 0.89 | 0.75 |
| | SIEMENS–ROCHE | 0.60 | 0.55 to 0.65 | 0.74 | 0.80 |
| | BECKMAN–ROCHE | 0.79 | 0.75 to 0.82 | 0.79 | 0.99 |
| **Serum Chloride** | BECKMAN–SIEMENS | 0.64 | 0.60 to 0.68 | 0.89 | 0.71 |
| **Serum Potassium** | BECKMAN-SIEMENS | 0.92 | 0.90 to 0.94 | 0.94 | 0.98 |
| **Serum Sodium** | BECKMAN–SIEMENS | 0.60 | 0.55 to 0.65 | 0.80 | 0.75 |
| **(E) Renal Function Markers** | | | | | |
| **Serum Creatinine** | BECKMAN-SIEMENS | 0.79 | 0.76 to 0.82 | 0.91 | 0.86 |
| | SIEMENS–ROCHE | 0.94 | 0.93 to 0.95 | 0.96 | 0.97 |
| | BECKMAN–ROCHE | 0.71 | 0.68 to 0.75 | 0.93 | 0.77 |
| **Blood Urea** | BECKMAN–SIEMENS | 0.97 | 0.96 to 0.97 | 0.99 | 0.98 |
| | SIEMENS–ROCHE | 0.98 | 0.97 to 0.98 | 0.99 | 0.98 |
| | BECKMAN–ROCHE | 0.98 | 0.98 to 0.98 | 0.98 | 0.99 |
| **Serum Uric acid** | BECKMAN–SIEMENS | 0.99 | 0.98 to 0.99 | 0.99 | 0.99 |
| | SIEMENS–ROCHE | 0.99 | 0.99 to 0.99 | 0.99 | 0.99 |
| | BECKMAN–ROCHE | 0.98 | 0.98 to 0.99 | 0.99 | 0.99 |

| | | | | | |
|---|---|---|---|---|---|
| **(F) HbA1c** | | | | | |
| **HbA1c** | Bio-Rad - Tosoh | 0.97 | 0.97 to 0.98 | 0.99 | 0.98 |
| **(G) Iron metabolisms** | | | | | |
| **UIBC** | BECKMAN-SIEMENS | 0.89 | 0.87 to 0.90 | 0.97 | 0.91 |
| | SIEMENS–ROCHE | 0.88 | 0.86 to 0.90 | 0.94 | 0.93 |
| | BECKMAN–ROCHE | 0.93 | 0.92 to 0.94 | 0.93 | 0.99 |
| **Serum Iron** | BECKMAN–SIEMENS | 0.98 | 0.98 to 0.98 | 0.98 | 0.99 |
| | SIEMENS–ROCHE | 0.94 | 0.93 to 0.95 | 0.95 | 0.99 |
| | BECKMAN–ROCHE | 0.94 | 0.93 to 0.95 | 0.95 | 0.99 |
| **Ferritin** | BECKMAN–SIEMENS | 0.95 | 0.94 to 0.95 | 0.99 | 0.96 |
| | SIEMENS–ROCHE | 0.88 | 0.86 to 0.89 | 0.99 | 0.88 |
| | BECKMAN–ROCHE | 0.74 | 0.72 to 0.76 | 0.98 | 0.75 |
| **(H) TSH** | | | | | |
| **TSH** | BECKMAN–SIEMENS | 0.98 | 0.97 to 0.98 | 0.98 | 0.99 |
| | SIEMENS–ROCHE | 0.96 | 0.95 to 0.97 | 0.96 | 0.99 |
| | BECKMAN–ROCHE | 0.96 | 0.96 to 0.97 | 0.97 | 0.99 |

**Table S3: Bland Altman Analysis results**

| Assay | Analyzers | Mean Difference | Lower limit of agreement | Upper limit of agreement |
|---|---|---|---|---|
| **(A)Liver function markers** | | | | |
| **AST/SGOT** | BECKMAN-SIEMENS | 0.06 | -3.53 | 3.66 |
| | SIEMENS–ROCHE | -2.71 | -8.25 | 2.82 |
| | BECKMAN–ROCHE | -2.65 | -8.30 | 2.99 |
| **ALT/SGPT** | BECKMAN-SIEMENS | -0.16 | -5.58 | 5.25 |
| | SIEMENS–ROCHE | -0.97 | -5.46 | 3.51 |
| | BECKMAN–ROCHE | -1.13 | -5.93 | 3.65 |
| **ALP** | BECKMAN–SIEMENS | 1.94 | -8.12 | 12.01 |
| | SIEMENS–ROCHE | -1.21 | -15.10 | 12.68 |
| | BECKMAN–ROCHE | 0.73 | -16.07 | 17.54 |
| **GGT** | BECKMAN–SIEMENS | 3.52 | 0.45 | 6.58 |
| | SIEMENS–ROCHE | -0.47 | -6.88 | 5.94 |
| | BECKMAN–ROCHE | 3.05 | -3.23 | 9.33 |
| **Serum Total Protein** | BECKMAN-SIEMENS | -0.07 | -0.36 | 0.20 |
| | SIEMENS–ROCHE | -0.13 | -0.44 | 0.17 |
| | BECKMAN–ROCHE | -0.21 | -0.54 | 0.11 |
| **Serum Albuin** | BECKMAN–SIEMENS | -0.04 | -0.31 | 0.22 |
| | SIEMENS–ROCHE | -0.07 | -0.29 | 0.13 |
| | BECKMAN–ROCHE | -0.12 | -0.33 | 0.08 |
| **Serum Bilirubin, (Total)** | BECKMAN–SIEMENS | 0.03 | -0.08 | 0.15 |
| | SIEMENS–ROCHE | 0.14 | -0.07 | 0.36 |
| | BECKMAN–ROCHE | 0.17 | -0.00 | 0.36 |
| **Serum Bilirubin, (Direct)** | BECKMAN–SIEMENS | -0.06 | -0.16 | 0.02 |
| | SIEMENS–ROCHE | -0.06 | -0.15 | 0.02 |
| | BECKMAN–ROCHE | -0.13 | -0.28 | 0.01 |

| | | | | |
|---|---|---|---|---|
| **(B) Lipid Parameters** | | | | |
| **Serum Total Cholesterol** | BECKMAN-SIEMENS | 2.18 | -6.48 | 10.85 |
| | SIEMENS–ROCHE | -6.03 | -17.16 | 5.09 |
| | BECKMAN–ROCHE | -3.84 | -11.59 | 3.89 |
| **Serum Triglycerides** | BECKMAN-SIEMENS | -9.72 | -17.87 | -1.57 |
| | SIEMENS–ROCHE | -3.47 | -15.61 | 8.66 |
| | BECKMAN–ROCHE | -13.19 | -25.80 | -0.59 |
| **Serum HDL Cholesterol** | BECKMAN–SIEMENS | 2.66 | -4.09 | 9.41 |
| | SIEMENS–ROCHE | 0.04 | -4.75 | 4.83 |
| | BECKMAN–ROCHE | 2.70 | -4.79 | 10.21 |
| **Serum LDL Cholesterol** | BECKMAN-SIEMENS | -7.24 | -29.55 | 15.06 |
| | SIEMENS–ROCHE | 6.70 | -5.56 | 18.98 |
| | BECKMAN–ROCHE | -0.53 | -18.1 | 17.1 |
| **Apolipoprotein A1** | BECKMAN–SIEMENS | 17.91 | 1.37 | 34.44 |
| **Apolipoprotein B** | BECKMAN–SIEMENS | 16.62 | 5.07 | 28.16 |
| **(C) Vitamins** | | | | |
| **Vitamin B12** | BECKMAN–SIEMENS | -154.7 | -281.71 | -27.83 |
| | SIEMENS–ROCHE | -4.84 | -238.96 | 229.28 |
| | BECKMAN–ROCHE | -159.61 | -409.93 | 90.70 |
| **VITAMIN D (25 OH VITAMIN D)** | BECKMAN–SIEMENS | -0.28 | -12.97 | 12.40 |
| | SIEMENS–ROCHE | 2.04 | -11.57 | 15.65 |
| | BECKMAN–ROCHE | 1.76 | -11.61 | 15.14 |

| | | | | |
|---|---|---|---|---|
| **(D) Electrolytes** | | | | |
| **Serum Phosphorus** | BECKMAN-SIEMENS | -0.05 | -0.27 | 0.17 |
| | SIEMENS–ROCHE | -0.20 | -0.80 | 0.40 |
| | BECKMAN–ROCHE | -0.25 | -0.83 | 0.33 |
| **Serum Calcium** | BECKMAN–SIEMENS | 0.31 | -0.03 | 0.67 |
| | SIEMENS–ROCHE | -0.27 | -0.83 | 0.28 |
| | BECKMAN–ROCHE | 0.04 | -0.45 | 0.53 |
| **Serum Chloride** | BECKMAN–SIEMENS | -2.21 | -4.45 | 0.01 |
| **Serum Potassium** | BECKMAN-SIEMENS | -0.08 | -0.40 | 0.23 |
| **Serum Sodium** | BECKMAN–SIEMENS | -1.86 | -4.80 | 1.08 |
| **(E) Renal Function Markers** | | | | |
| **Serum Creatinine** | BECKMAN-SIEMENS | -0.09 | -0.24 | 0.04 |
| | SIEMENS–ROCHE | -0.04 | -0.13 | 0.05 |
| | BECKMAN–ROCHE | -0.14 | -0.27 | -0.001 |
| **Blood Urea** | BECKMAN–SIEMENS | -1.57 | -3.84 | 0.68 |
| | SIEMENS–ROCHE | 1.24 | -0.80 | 3.28 |
| | BECKMAN–ROCHE | -0.33 | -3.01 | 2.34 |
| **Serum Uric acid** | BECKMAN–SIEMENS | -0.03 | -0.38 | 0.31 |
| | SIEMENS–ROCHE | 0.08 | -0.16 | 0.33 |
| | BECKMAN–ROCHE | 0.05 | -0.31 | 0.416 |
| **(F) HbA1c** | | | | |
| **HbA1c** | Bio-Rad - Tosoh | -0.18 | -0.39 | 0.03 |

| | | | | |
|---|---|---|---|---|
| **(G) Iron metabolisms** | | | | |
| **UIBC** | BECKMAN-SIEMENS | -26.42 | -62.43 | 9.59 |
| | SIEMENS–ROCHE | 24.13 | -23.07 | 71.34 |
| | BECKMAN–ROCHE | -2.29 | -46.87 | 42.29 |
| **Serum Iron** | BECKMAN–SIEMENS | -0.22 | -10.31 | 9.86 |
| | SIEMENS–ROCHE | -3.61 | -22.19 | 14.97 |
| | BECKMAN–ROCHE | -3.83 | -22.13 | 14.47 |
| **Ferritin** | BECKMAN–SIEMENS | -12.55 | -42.90 | 17.79 |
| | SIEMENS–ROCHE | -30.31 | -92.13 | 31.50 |
| | BECKMAN–ROCHE | -42.86 | -131.77 | 46.04 |
| **(H) TSH** | | | | |
| **TSH** | BECKMAN–SIEMENS | -0.08 | -0.77 | 0.61 |
| | SIEMENS–ROCHE | -0.07 | -1.09 | 0.93 |
| | BECKMAN–ROCHE | -0.16 | -1.04 | 0.72 |

**Abbreviations:**

1. AST/SGOT - Aspartate Aminotransferase
2. ALT/SGPT - Alanine Aminotransferase
3. ALP - Alkaline Phosphatase
4. GGT - Gamma-Glutamyl Transferase
5. HbA1c - Glycosylated Haemoglobin
6. UIBC - Unsaturated Iron Binding Capacity
7. TSH - Thyroid-Stimulating Hormone

### Supplementary Figures:

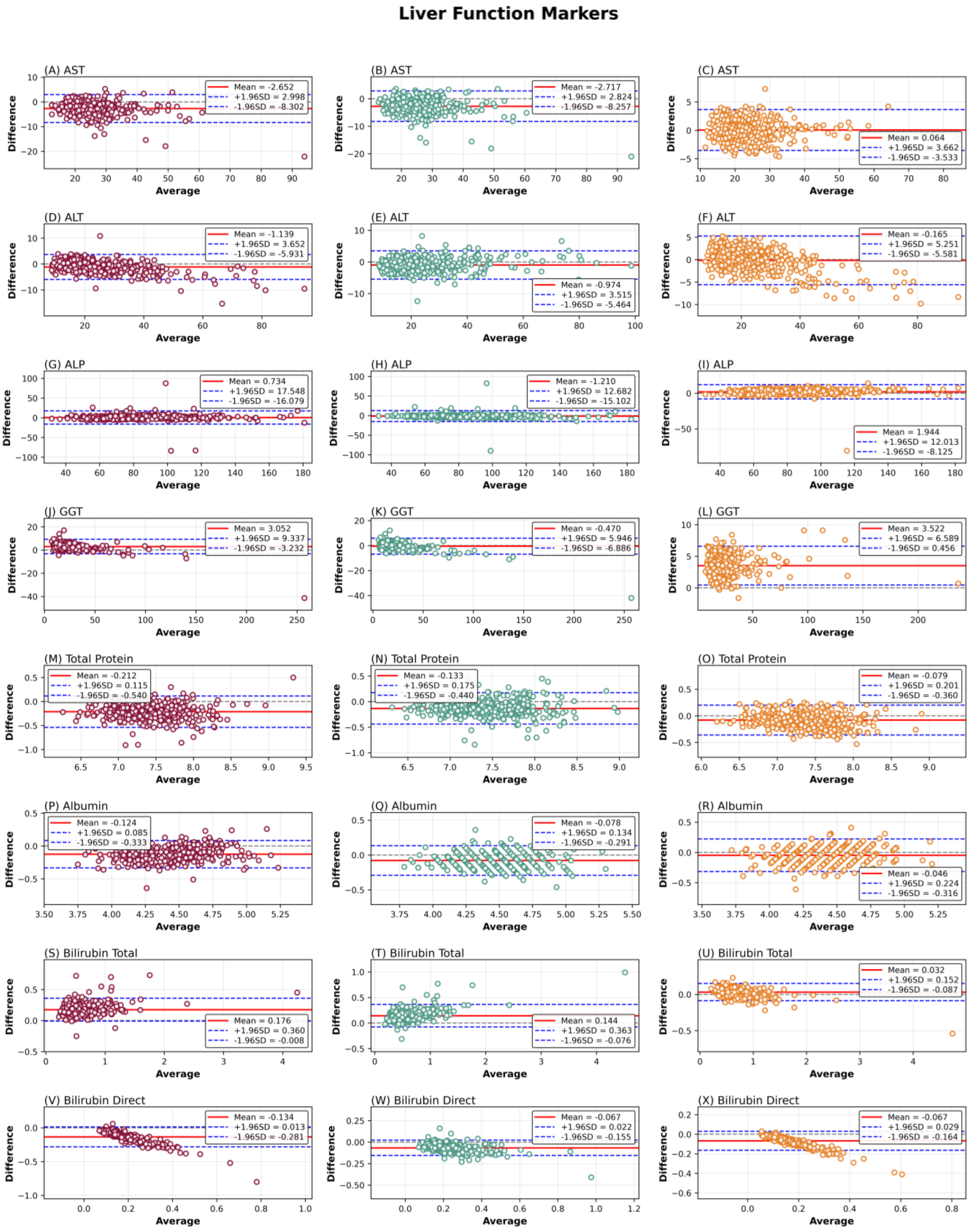

**Figure S1:** Bland-Altman analysis of agreement between clinical chemistry analyzers for liver function markers. Bland-Altman plots display the mean bias and 95% limits of agreement for AST(A-C), ALT (D-F), ALP(G-I), GGT(J-L), Total protein (M-O), Albumin(P-R), Bilirubin total (S-U), and Bilirubin direct (V-X) measured across different analyzers. The solid horizontal line represents the mean difference (bias), while the dashed lines indicate the upper and lower limits of agreement (mean ± 1.96 SD). Comparisons are shown for three analyzer pairings: Beckman-Roche (maroon), Siemens-Roche (green) and Beckman-Siemens (orange).

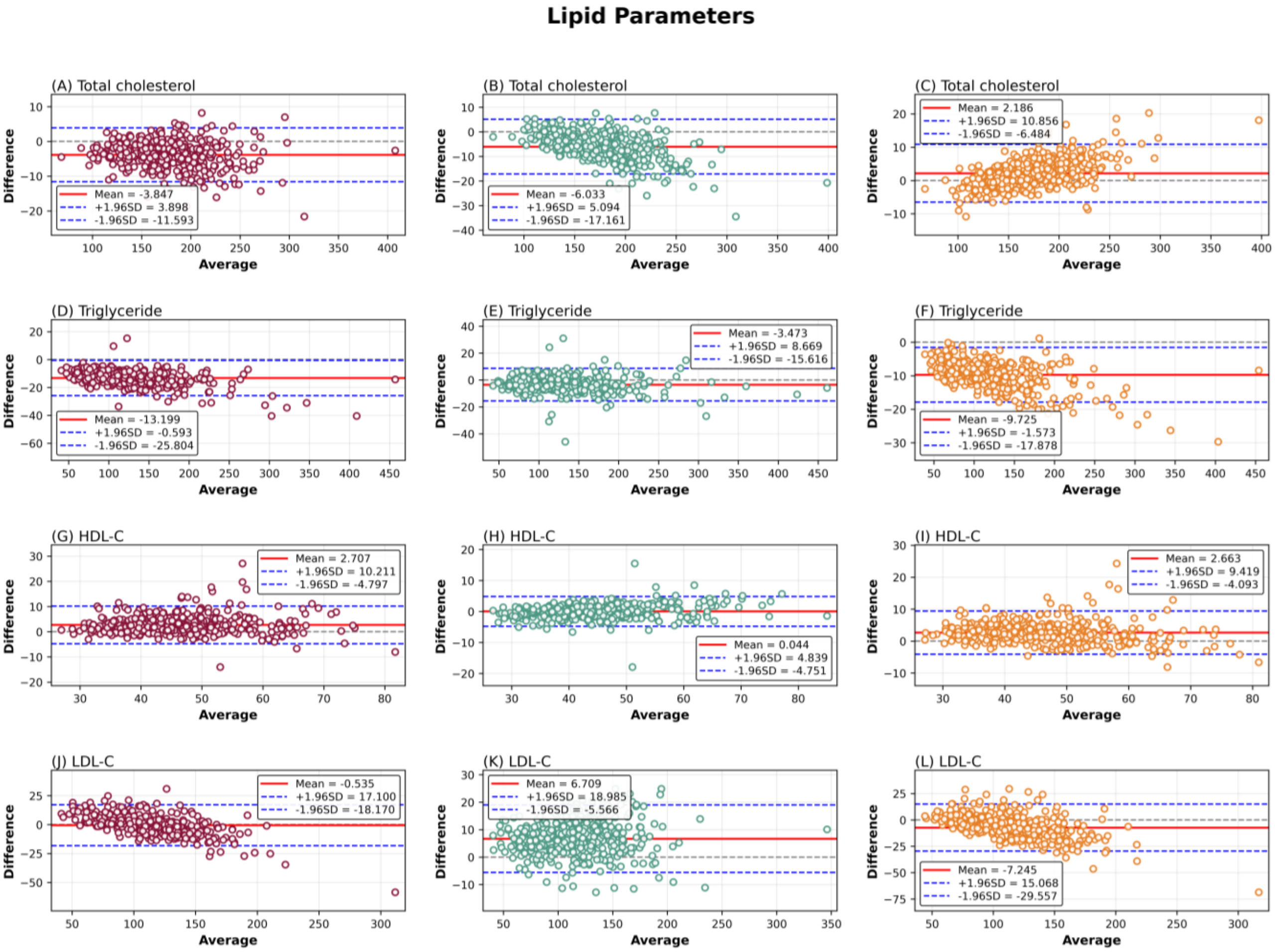

**Figure S2:** Bland-Altman analysis of agreement between clinical chemistry analyzers for lipid parameters. Bland-Altman plots illustrate the mean bias and 95% limits of agreement for Total cholesterol (A-C), Triglycerides (D-F), HDL-C (G-I) and LDL-C (J-L). The central horizontal line denotes the mean difference, and dashed lines represent the 95% limits of agreement (mean ± 1.96 SD). Three analyzer pairings are shown: Beckman-Roche (maroon), Siemens-Roche (green) and Beckman-Siemens (orange).

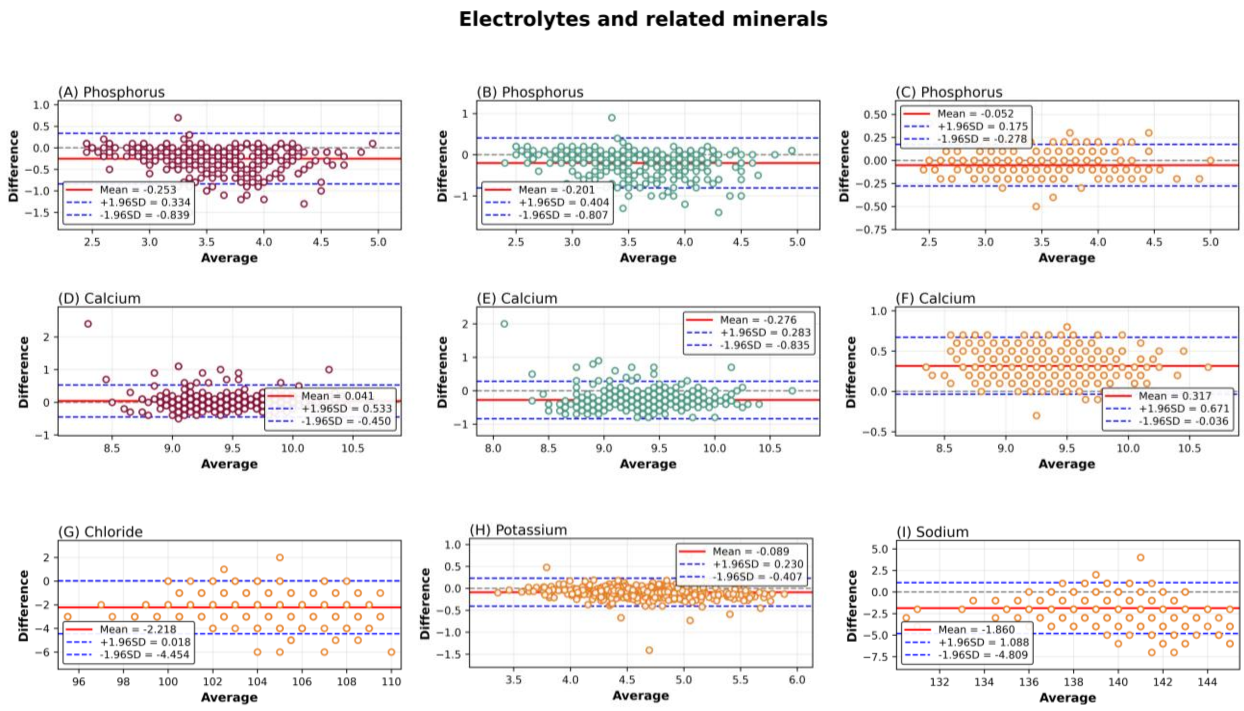

**Figure S3:** Bland-Altman analysis of agreement between clinical chemistry analyzers for electrolytes and related minerals. Bland-Altman plots depict the mean bias and 95% limits of agreement for Phosphorus (A-C), Calcium (D-F), Chloride (G), Potassium, (H) and Sodium (I) across different analyzers. The mean difference is indicated by the solid horizontal line, while dashed lines represent the upper and lower limits of agreement (mean ± 1.96 SD). Beckman-Roche (maroon), Siemens-Roche (green) and Beckman-Siemens (orange).

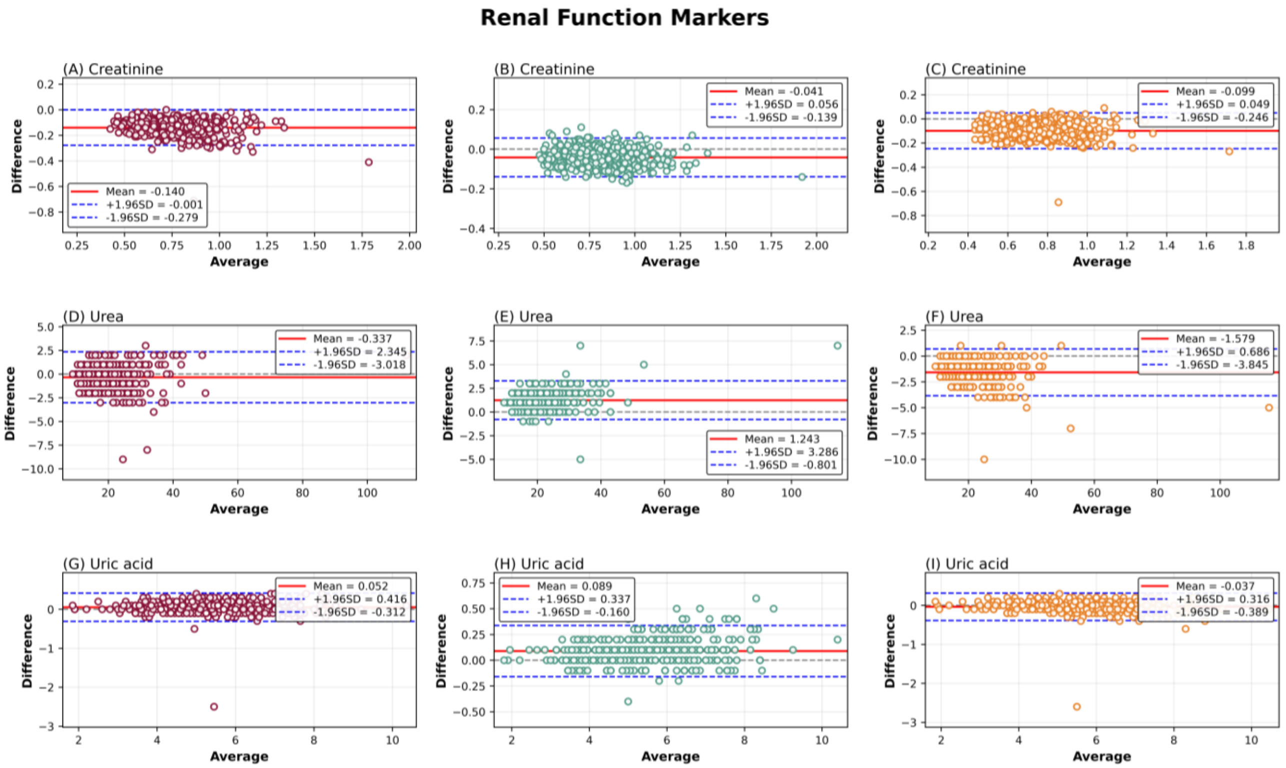

**Figure S4:** Bland-Altman analysis of agreement between clinical chemistry analyzers for renal function markers. Bland-Altman plots show the mean bias and 95% limits of agreement for Creatinine (A-C), Urea (D-F), and Uric acid (G-I) measured across analyzers. The solid horizontal line represents the mean difference, and the dashed lines correspond to the limits of agreement (mean ± 1.96 SD). Analyzer comparisons include Beckman-Roche (maroon), Siemens-Roche (green) and Beckman-Siemens (orange).

### Thyroid Stimulating Hormone

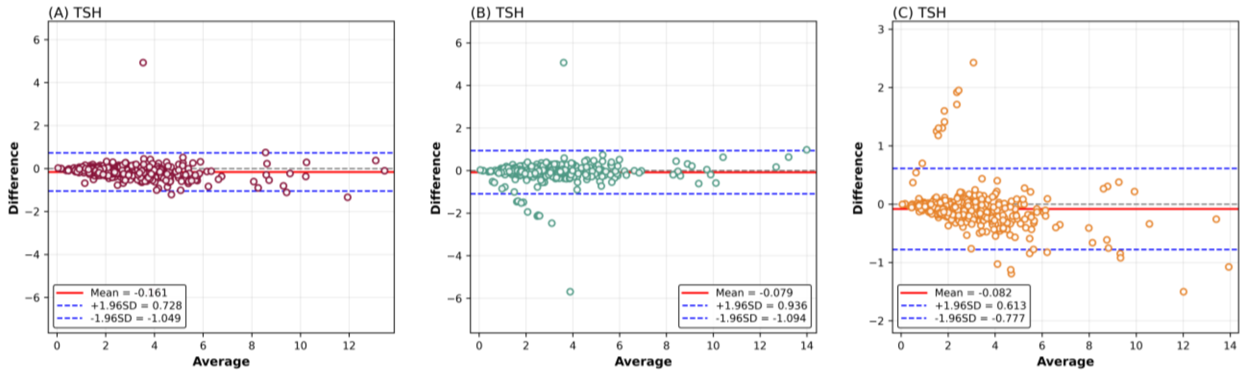

**Figure S5:** Bland-Altman analysis of agreement between immunoassay analyzers for TSH. Bland-Altman plots display the mean bias and 95% limits of agreement for Iron measurements across the compared analyzers. The solid horizontal line indicates the mean difference, while dashed lines denote the upper and lower limits of agreement (mean ± 1.96 SD). Analyzer comparisons include Beckman-Roche (maroon), Siemens-Roche (green) and Beckman-Siemens (orange).

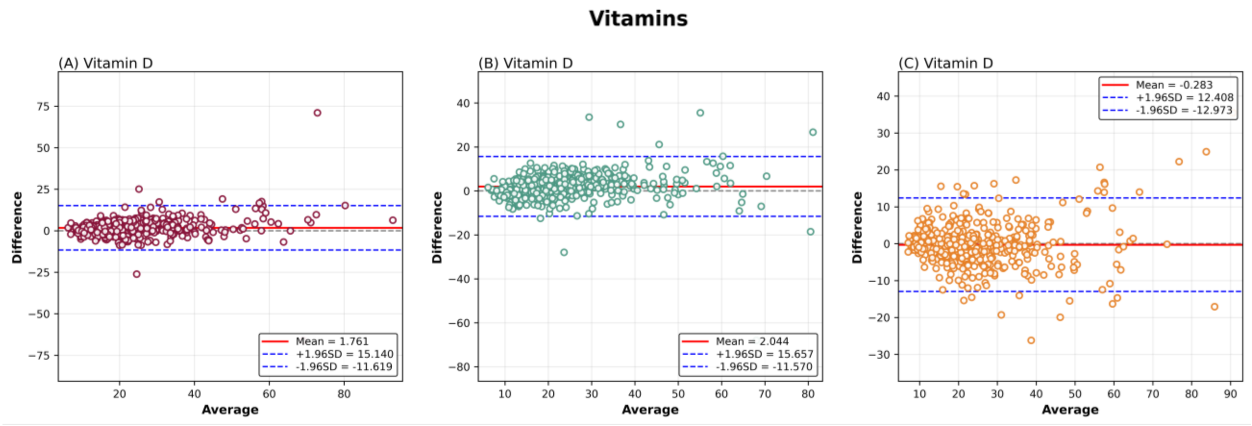

**Figure S6:** Bland-Altman analysis of agreement between immunoassay analyzers for vitamin measurements. Bland-Altman plots display the mean bias and 95% limits of agreement for Vitamin D concentrations across the compared analyzers. The solid horizontal line represents the mean difference, while dashed lines indicate the upper and lower limits of agreement (mean ± 1.96 SD). Analyzer comparisons include Beckman-Roche (maroon), Siemens-Roche (green) and Beckman-Siemens (orange).

### HbA1c

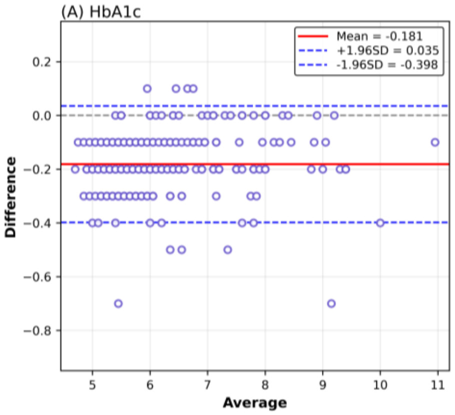

**Figure S7:** Bland-Altman analysis of agreement between analyzers for glycated haemoglobin (HbA1c). Bland-Altman plots present the mean bias and 95% limits of agreement for HbA1c values measured across different platforms. The solid line indicates the mean difference, and dashed lines represent the limits of agreement (mean ± 1.96 SD). Analyzer comparisons include Bio-rad - Tosoh.
